# Effectiveness of the WHO-authorized Covid-19 Vaccines: a Rapid Review of Global Reports till June 30, 2021

**DOI:** 10.1101/2021.08.23.21262500

**Authors:** Chang-Jie Cheng, Chun-Yi Lu, Ya-Hui Chang, Yu Sun, Hai-Jui Chu, Chun-Yu Lee, Chang-Hsiu Liu, Cheng-Huai Lin, Chien-Jung Lu, Chung-Yi Li

## Abstract

**Objective:** Large clinical trials have proved the efficacy of Covid-19 vaccine, and the number of literature about the effectiveness is rapidly growing in the first half of year after mass vaccination was administrated globally. This rapid review aims to provide evidence syntheses as a means to complement the current evidence on the vaccine effectiveness (VE) against various outcomes in real-world settings.

**Methods:** This review is conducted based on the updated guideline of PRISMA 2020. Databases (PubMed, EMBASE, and MedRxiv) were searched up to 30 June 2021, (PROSPERO ID: 266866). The studies that assessed the VE of the 6 WHO-authorized vaccines (BNT162b2, ChAdOx1, Ad26.COV2.S, mRNA-1273, BBIBP-CorV, and CoronaVac) were eligible to be included. Quality assessment was performed based on ROBINS-I by 2 independent reviewers.

**Findings:** A total of 39 studies were included, covering over 15 million of participants from 11 nations. Among the general population after 2 doses of vaccination, the VE against symptomatic SARS-CoV-2 infection was estimated at 89%–97%, 92% (95% CI, 78%–97%) and 94% (95% CI, 86%–97%) for BNT162b2, ChAdOx1 and mRNA-1273, respectively. As for the protective effects against B.1.617.2 related symptomatic infection, the VE was 88% (95% CI, 85.3%–90.1%) by BNT162b2 and 67.0% (95% CI, 61.3%–71.8%) by ChAdOx1 after fully vaccination.

**Conclusion:** This review revealed a consistently high effectiveness of vaccines among the general population in real-world settings. Further studies are needed to provide the information on different races/ethnicity, the effects against SARS-CoV-2 variants, and the duration of protection with longer study time.

## Introduction

Large randomized control trials have demonstrated efficacy against SARS-CoV-2 infection to be 70%, 94%, 95%, and 78% after two doses of ChAdOx1 (Oxford/AstraZeneca), mRNA-1273 (Moderna), BNT162b2 (BioNTech/Pfizer), and BBIBP-CorV (Sinopharm) vaccines, respectively.^1–4^ Single-dose of Ad26.COV2.S (Johnson & Johnson) vaccine yielded an efficacy of 81.7% against severe-critical Covid-19 disease.^5^ Based on the good results from clinical trials, Covid-19 vaccination programs have been extensively rolled out in many countries around the world. But over the first half-year since vaccine administration globally, most countries have fully vaccination rates less than 50%.^6^ Billions of people in the world are eagerly waiting for the Covid-19 vaccines on one hand, and questioning about how well the vaccines work in the real world on the other hand. As relevant study reports being released successively, a wide range of vaccine effectiveness (VE) has been noticed. The estimated effectiveness after full vaccination could range from 50%^7^ to 100%,^8^ according to effectiveness studies of the 6 vaccines (BNT162b2, ChAdOx1, Ad26.COV2.S, mRNA-1273, BBIBP-CorV, CoronaVac [Sinovac]) which were listed on the World Health Organization (WHO) Emergency Use Listing as of June 2021.^9^

Given different efficacy was shown for vaccines developed with different platforms,^1–4^ their effectiveness in the real world for different populations needs to be confirmed. A Denmark study showed different VE in different age groups (77% for people ≥85 years of age vs. 86% for people ≥65 years), and different living/working environments (53% for long-term care facilities dwellers vs. 80% for healthcare workers).^10^ Similarly, the VE may also be different among different countries or races. Attentions should also be paid when VE from different studies are compared as what outcomes were chosen in determining VE. A study on the residents of long-term care facilities in Spain showed the VE of mRNA vaccines was 70% against asymptomatic infection and 97% against death.^11^ In addition, many SARS-CoV-2 variants have been evolving since the pandemic. B.1.1.7 (alpha), B.1.351 (beta), B.1.617.2 (delta), and P.1 (gamma) variants which are circulating in the whole world and are causing serious infections and mortality rate are classified as variants of concern (VOC) by WHO. The VE against these VOC needs to be explored from the literature.

This rapid review aims to assess the effectiveness of WHO-authorized Covid-19 vaccines, taking into account of aforementioned factors including country, characteristics of study population, study design, outcomes, and the analysis of involved VOC.

## Methods

### Search strategy and selection criteria

This rapid review is conducted based on the updated guideline of PRISMA 2020 statement and its recommended checklist.^12, 13^ We have registered this review on PROSPERO (ID: 266866) on July 13, 2021. The eligibility criteria are reports evaluating the effectiveness of Covid-19 vaccines in populations aged ≥16 years. Additional inclusion criteria require that the sample size of the vaccinated population should be more than 1,000 to have sufficient event number. Two authors (CJC, YS) did the literature search from PubMed, EMBASE and medRxiv. The search terms in PubMed and EMBASE were “effectiveness”, “Covid-19 vaccine”, and publish time “2021”. Preprint articles from medRxiv were searched with the terms “effectiveness Covid-19 vaccine” or “effectiveness SARS-CoV-2 vaccine” in the titles or abstracts.

The article types we reviewed included original investigation, research letter, short communication, and correspondence articles. While screening the title and abstracts of the relevant articles in PubMed, similar researches with titles shown on the web page were also checked. We updated our search up to June 30, 2021. CJC and YS contributed to the titles and abstracts screening for relevance and reviewing of full-text articles against inclusion and exclusion criteria. We excluded the following researches: in vitro studies, animal studies, experimental clinical trials, systematic reviews or meta-analyses, diagnostic studies, methodological publications, editorial-style reviews, abstracts of posters, secondary analyses, studies with only immunogenicity data, safety reports or post-infection treatment, and articles with analyses only on very specific target population such as veterans, dentists, pregnant women, and patients with malignancy or mental illness.

### Quality assessment, data extraction and synthesis

After identification of all relevant articles, quality assessment was performed based on ROBINS-I of Cochrane Handbook to assess the risk of bias. Each bias domain and overall risk of bias will be judged as “Low”, “Moderate”, “Serious”, or “Critical” risk of bias based on the check list on the ROBINS-I assessment chart. The extracted data included the following items: author, country, number of vaccinated and unvaccinated participants, study design, age and characteristic of participants, types of vaccine, outcomes, definition about minimal intervals between vaccination (first dose and second dose), and event measurement, involvement of SARS-CoV-2 VOC, and VE with confidence interval (CI) respectively after first dose and second dose of vaccine.

The formula for calculating VE is (1 - hazard ratio for SARS-CoV-2 infection in vaccinated vs. unvaccinated participants) × 100%. In studies that reported the incidence of infection, we calculated the incidence rate ratio (IRR) and convert it to unadjusted VE as (1- IRR in vaccinated vs. unvaccinated participants) × 100%. The VE from a case-control study is calculated as (1 - odds ratio) × 100%. In case of insufficient data in an article, we contacted the authors to obtain the required information by email. We applied narrative synthesis to process the data from included studies. As the number of literatures of VE is constantly growing over our processing period, this review only included reports which are released before July 1, 2021. Distributions of the VE estimates derived from the included studies were further graphically presented by Box plots, according to study population, brand of vaccine, variant, number of dose, and outcome.

## Results

### Study selection and characteristics

Of 2369 searched articles (2085 from PubMed, 195 from EMBASE, and 89 preprints from MedRxiv), 2312 were excluded while screening on abstracts and titles. After a full-text review on the remaining 57 articles for eligibility, 39 studies ^8, 10, 11, 14–49^ that met the inclusion criteria were included in this rapid review (Fig. 1, Supplementary Table S1). Results of quality assessment of all the included studies were shown in Supplementary Table S2. Before July 1, 2021, 24 of 39 studies are published in peer-reviewed journals ^8, 11, 14–16, 18, 21, 23, 24, 27–30, 32, 33, 35, 36, 39–41, 46, 47, 49, 50^ while the rest 15 studies are posted online as preprint articles.^10, 17, 19, 20, 22, 25, 26, 31, 34, 37, 38, 42, 43, 45, 48^ The characteristics of studies were shown in Supplementary Table S1, including country, study design, types of vaccines, outcomes, and SARS-CoV-2 variants involved in studies. The outcomes include all laboratory-confirmed SARS-CoV-2 infection, asymptomatic and symptomatic infection, hospitalization, critical disease, and death. As for evaluating the protective effects of the vaccine on SARS-CoV-2 variants, 5 studies mentioned the approximate prevalence of variants in the region of study population,^17, 18, 27, 33, 48^ and 11 studies calculated the number or percentage of cases with variants among all or sampled participants with a positive test of SARS-CoV-2 variants.^19, 25, 30, 32, 35–37, 41, 42, 45, 47^ Only 3 studies evaluated the VE against specific VOC, in which one study from Qatar reported the VE against B.1.1.7 and B.1.351,^8^ one research from Canada studied VE against B.1.1.7 and P.1,^45^ and the other one study from the UK evaluated the VE against B.1.1.7 and B.1.617.2 (delta variant).^19^

**Fig. 1.**
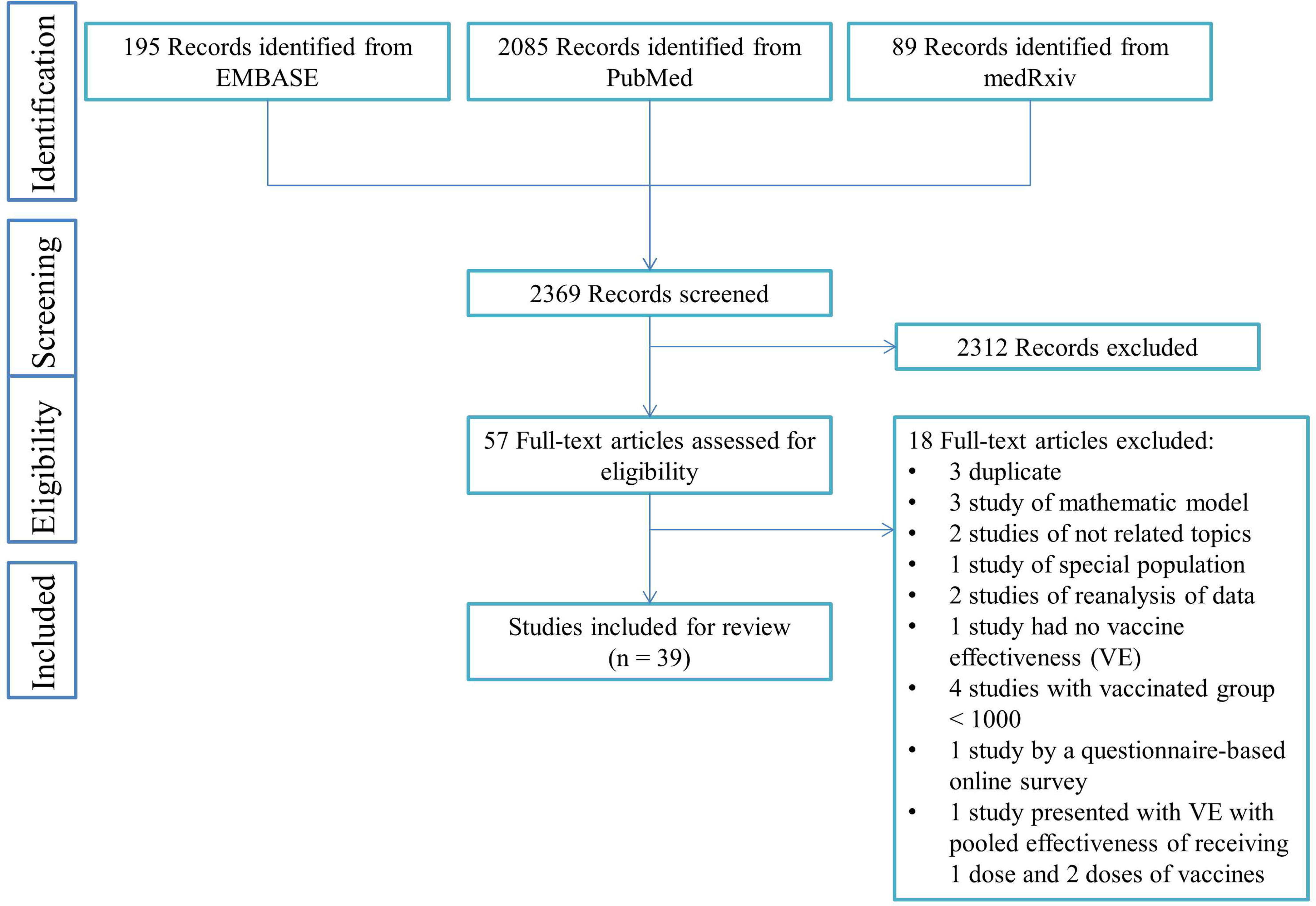
Flowchart of the literature searches

**Table 1.**
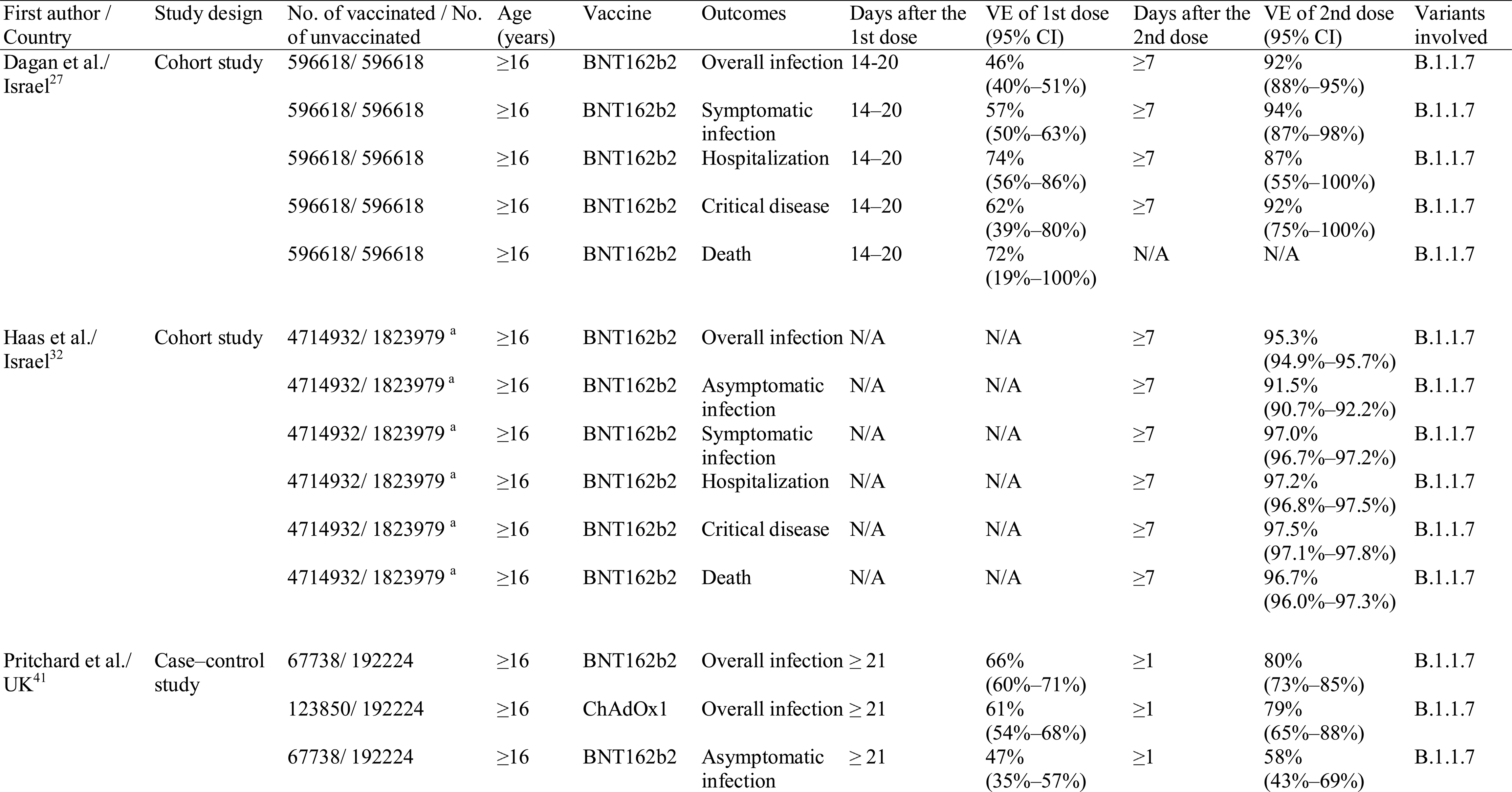
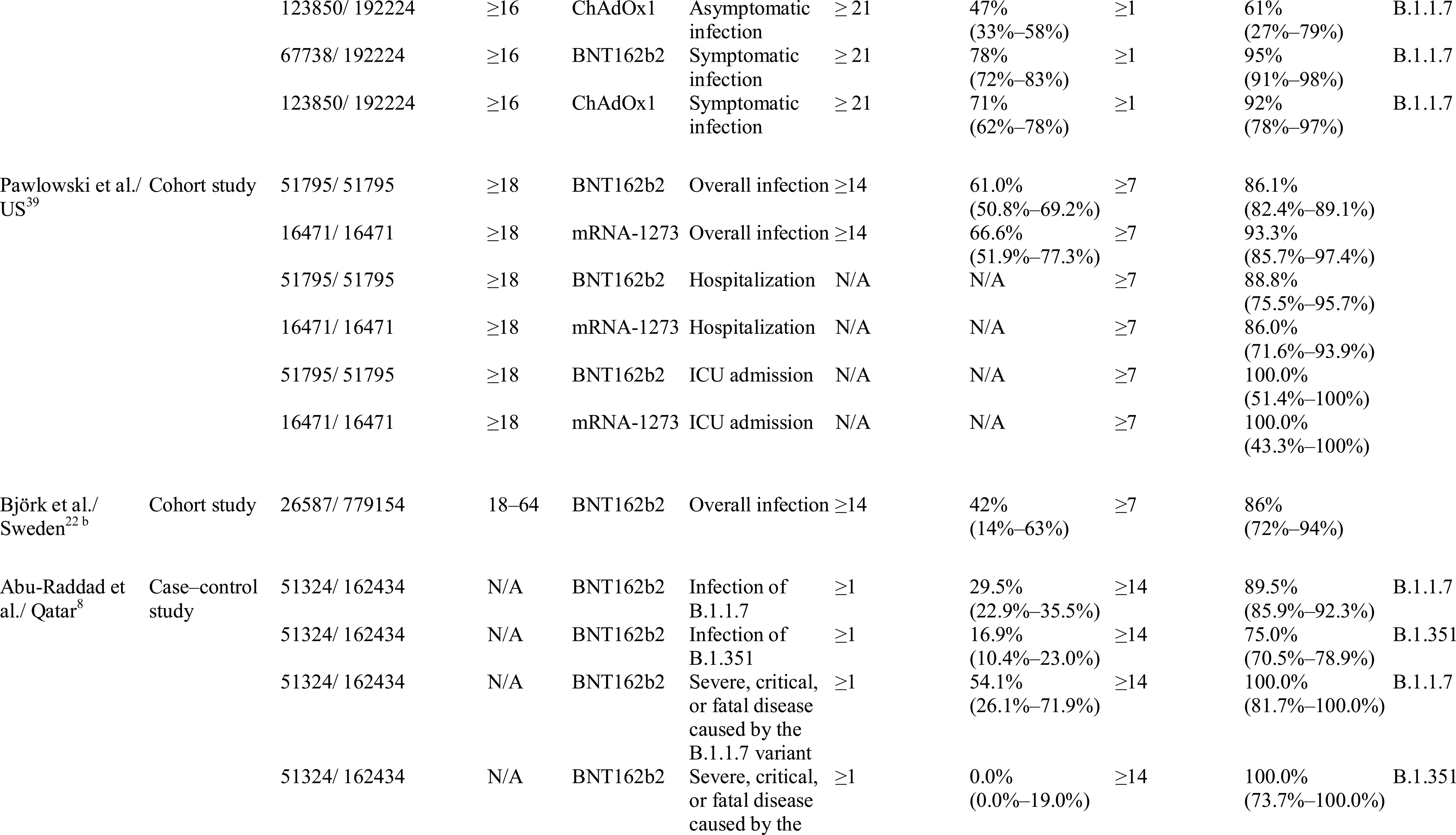
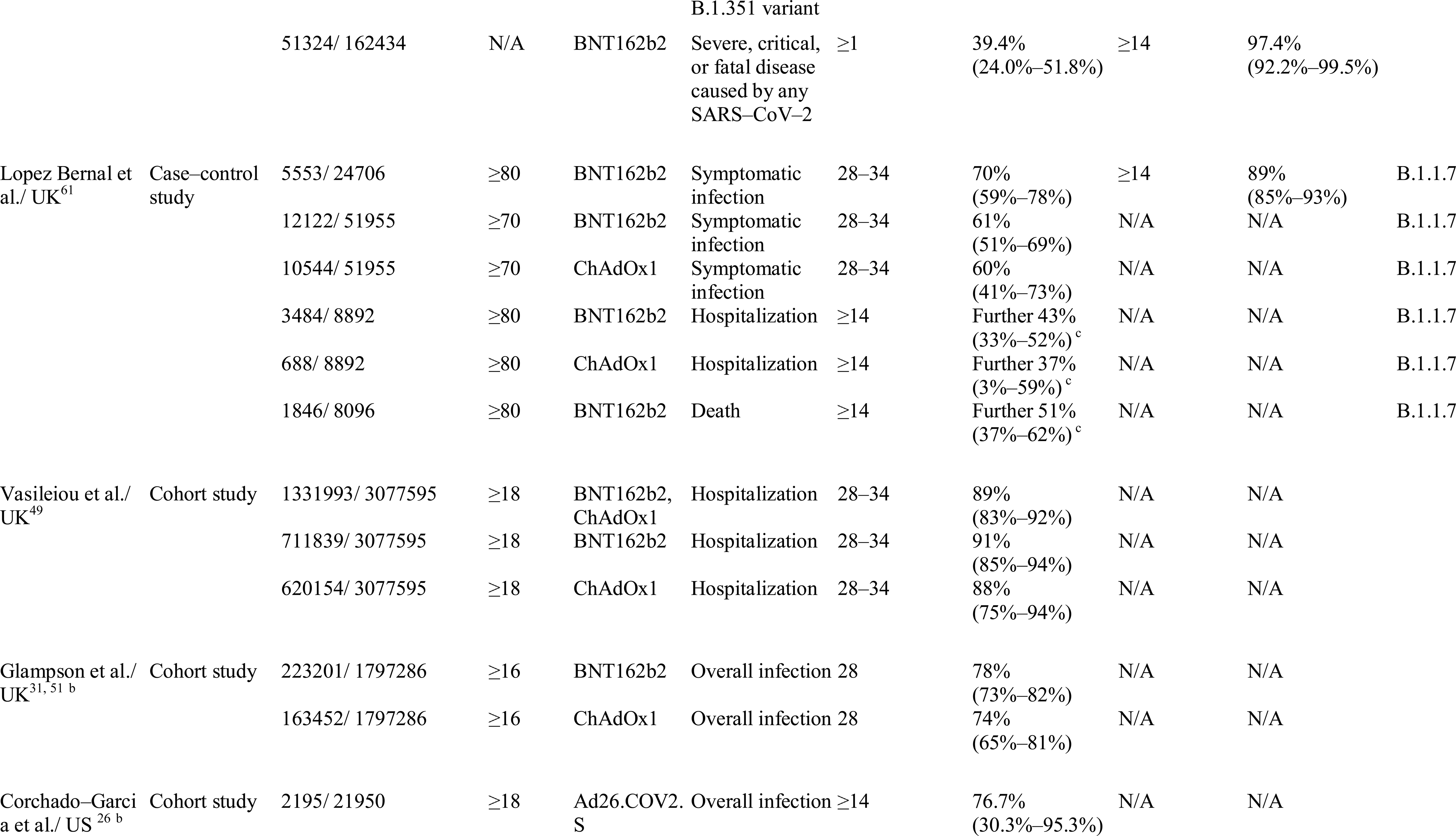
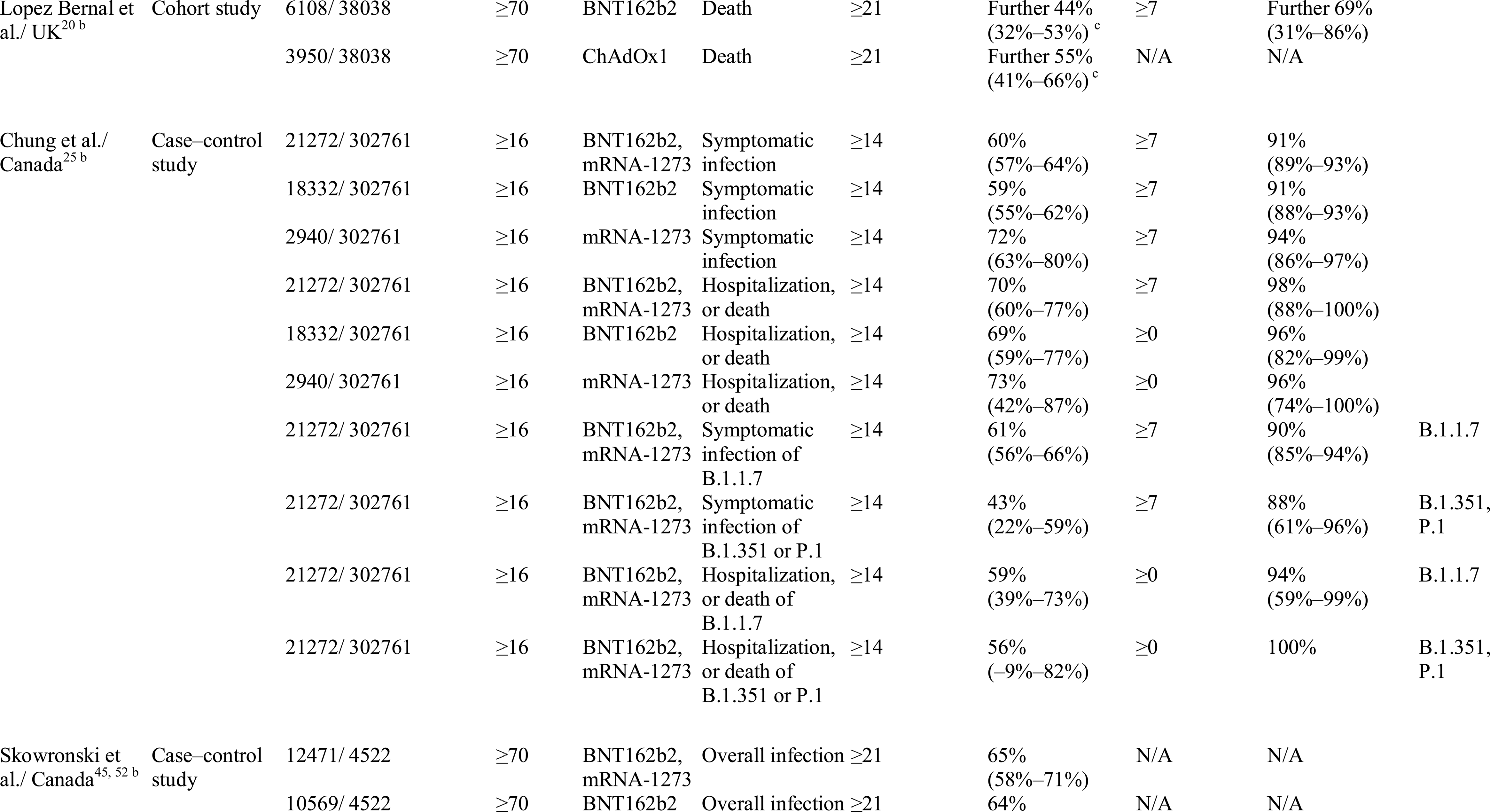
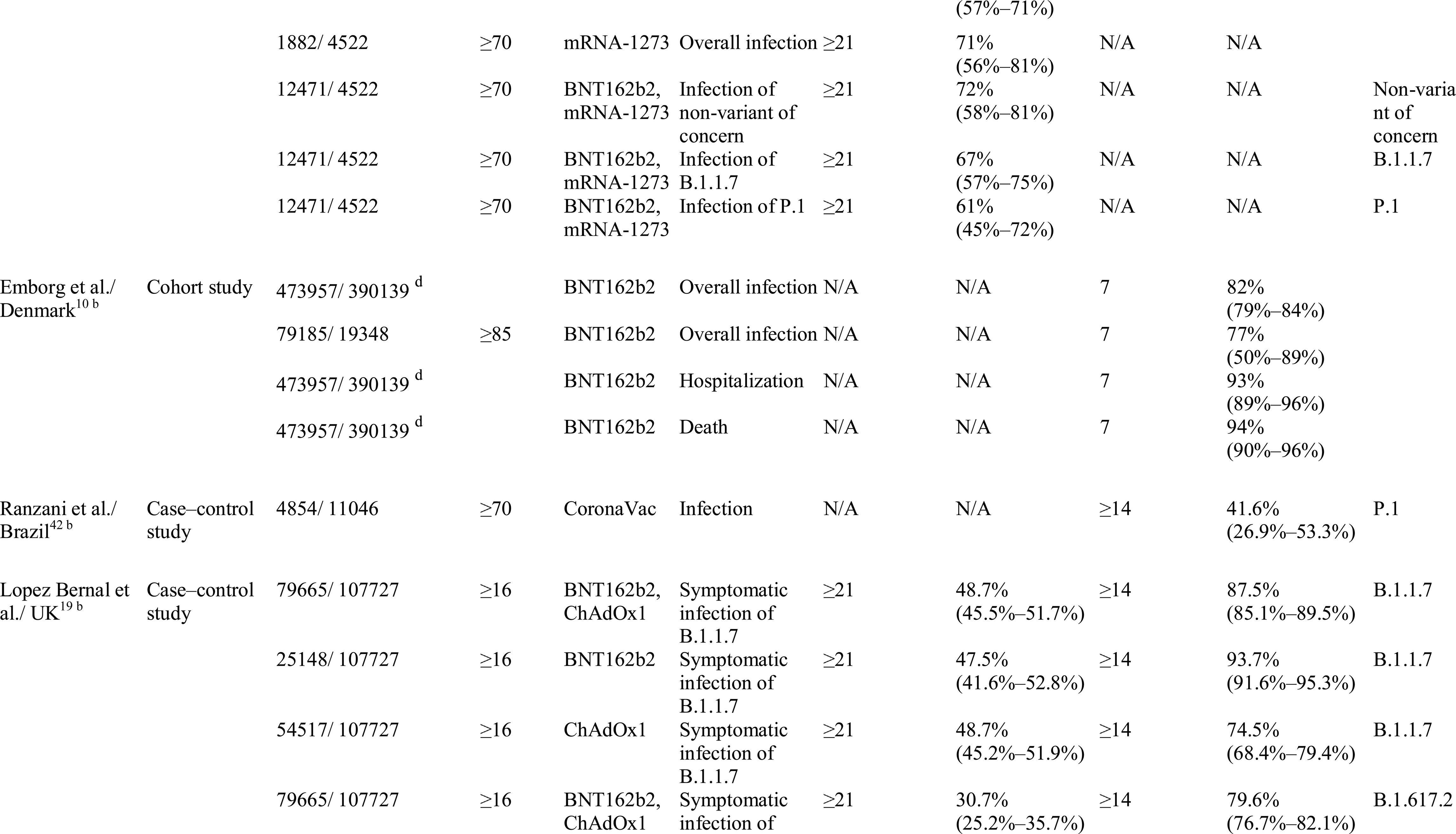
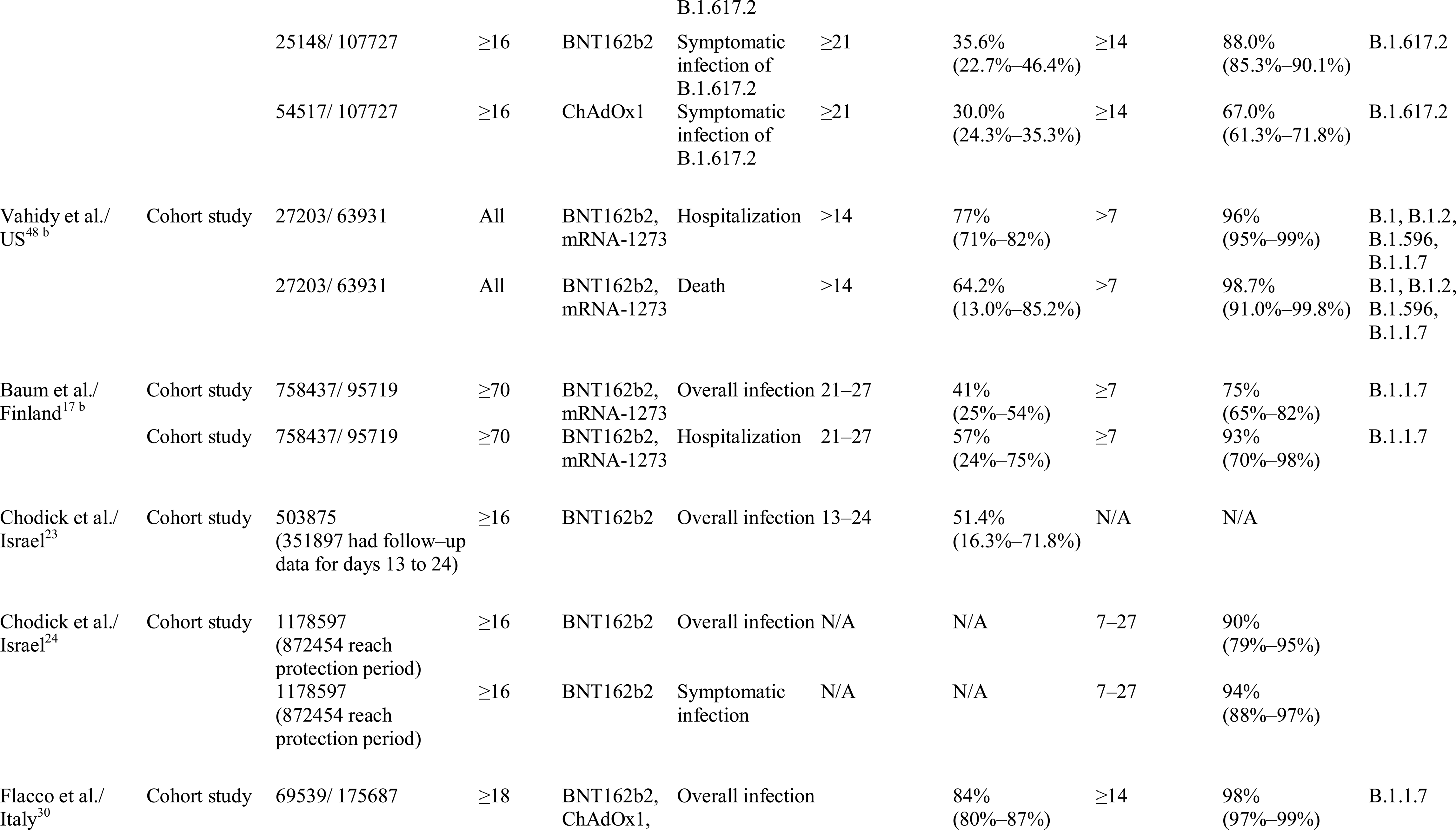
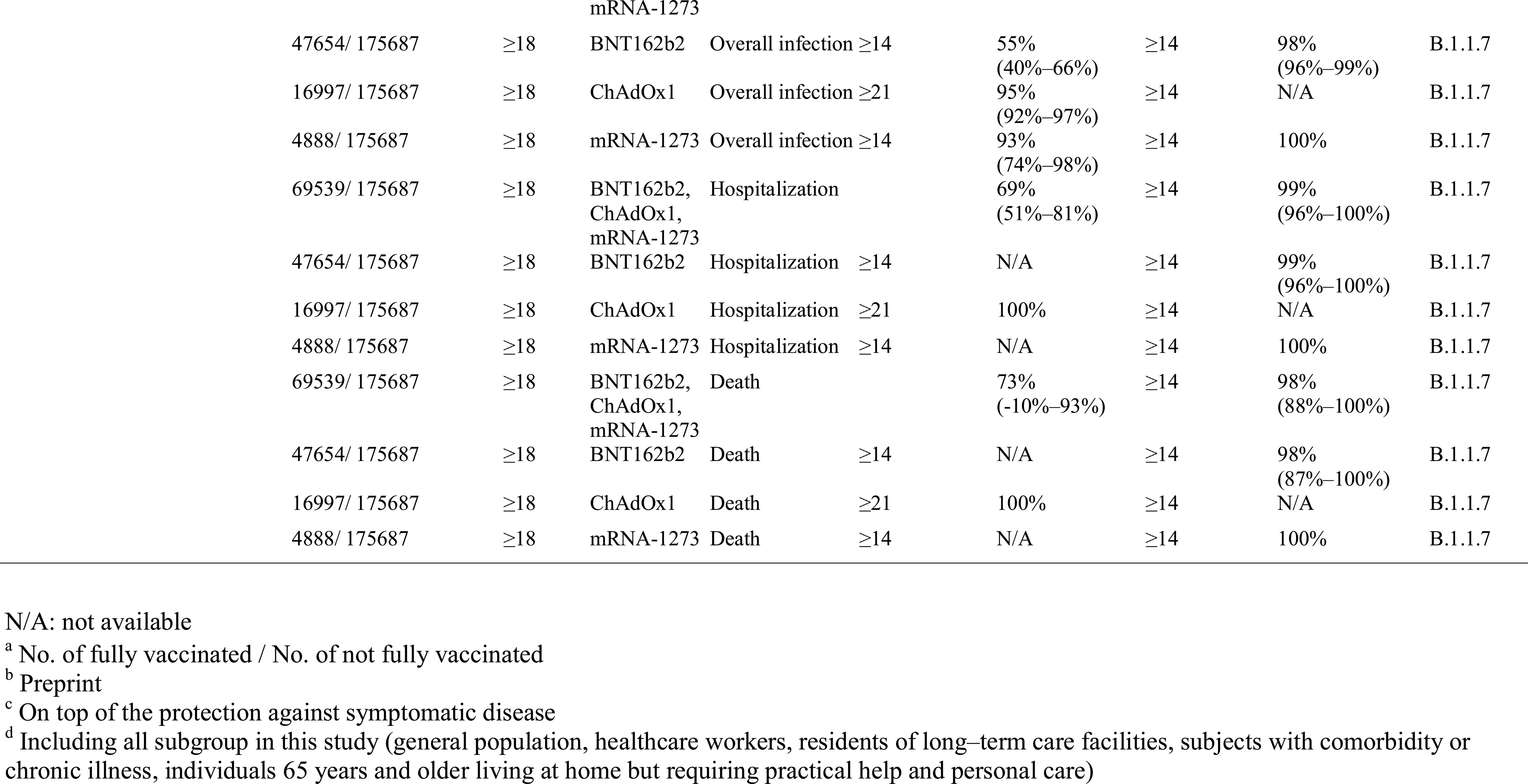
Summary of the studies on the effectiveness of COVID-19 vaccines among general population

**Table 2.**
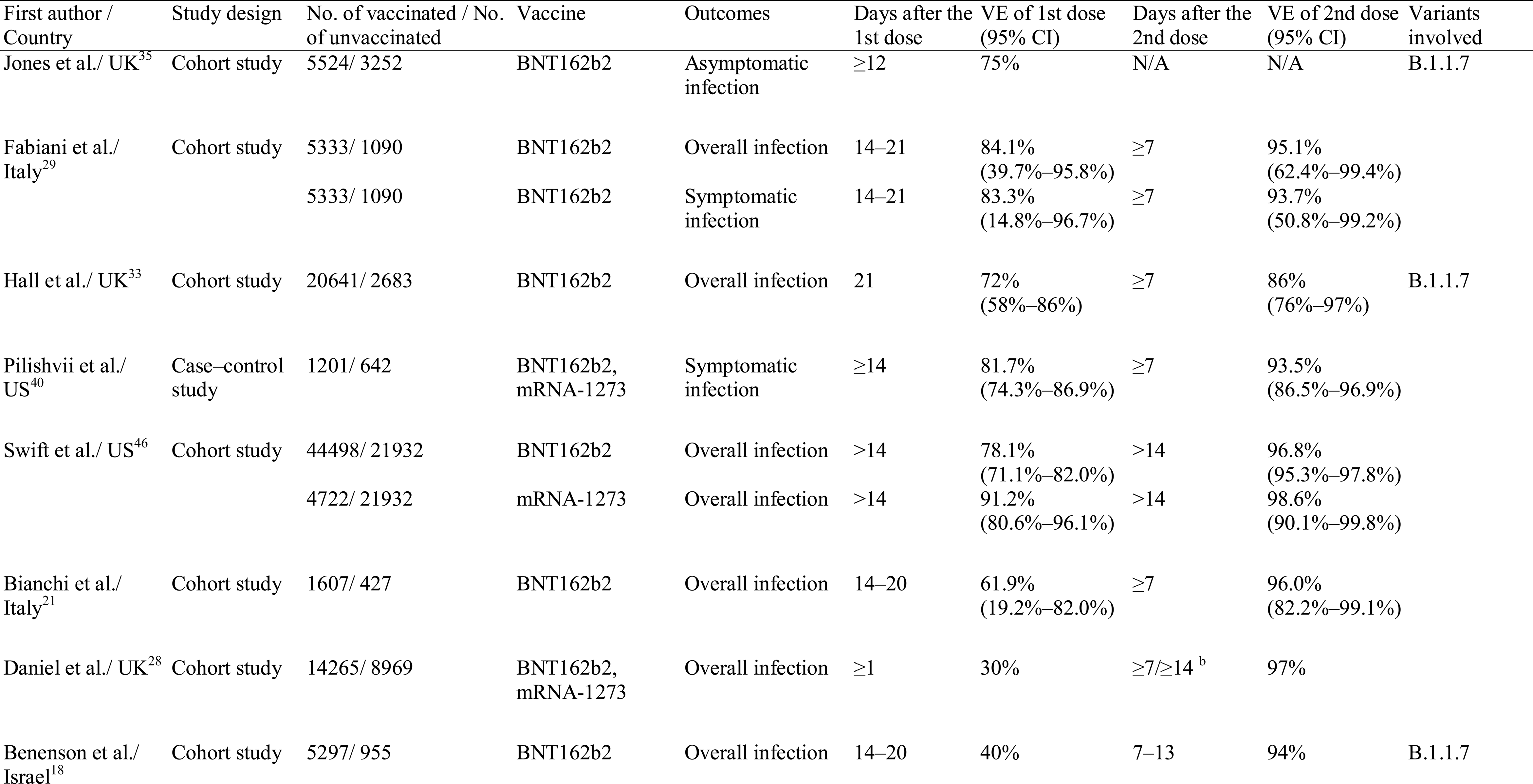
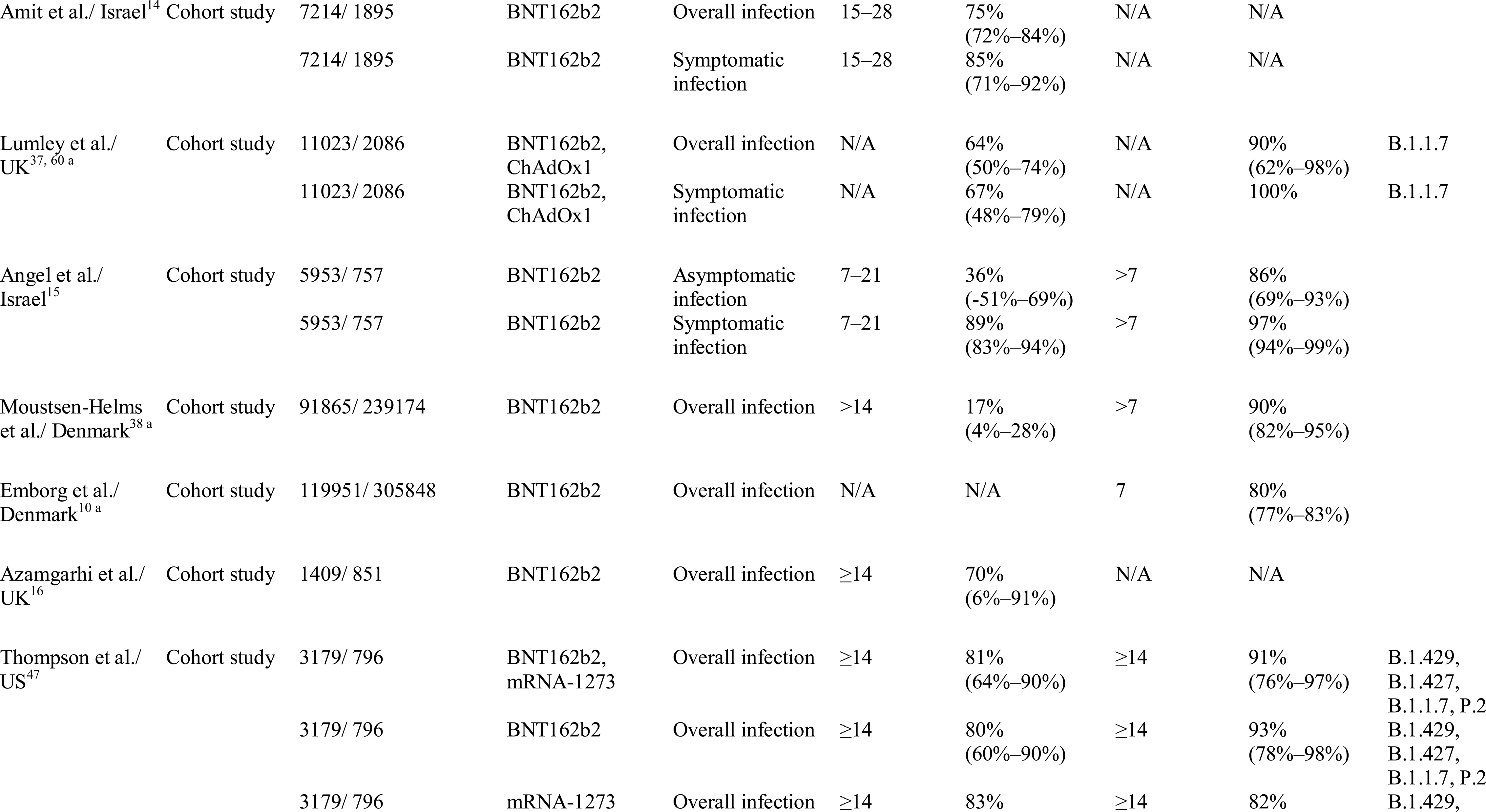
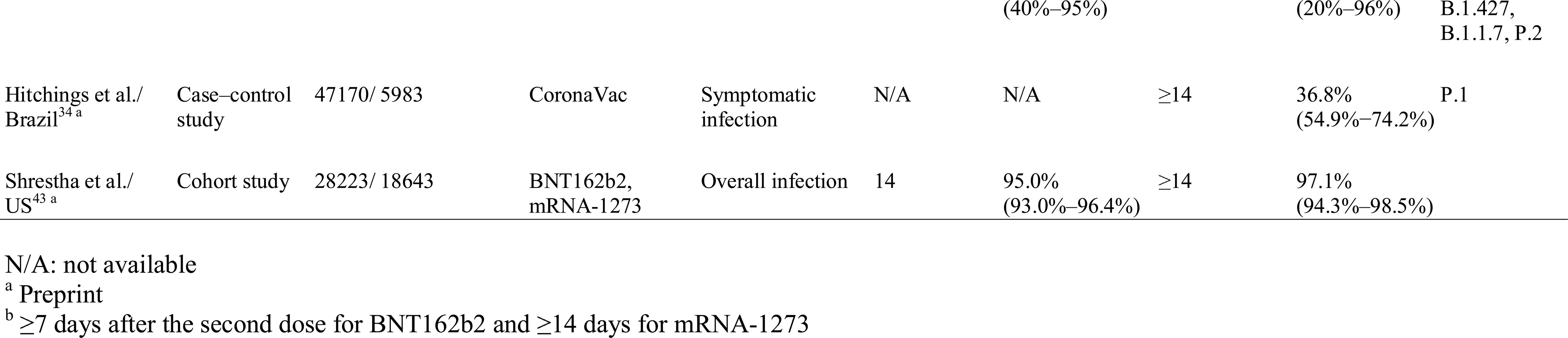
Summary of the studies on the effectiveness of COVID-19 vaccines among healthcare workers

### Effectiveness after partial or fully vaccinated for various outcomes

#### Overall SARS-CoV-2 infection across general and sub-populations

The characteristics of studies and their VE estimates against overall SARS-CoV-2 infection among general and specific populations are shown in Table 1–3 and Fig. 2. Among partially vaccinated general population, the VE varied among studies (42%–78% by BNT162b2,^22, 27, 30, 39, 41, 51, 52^ 67%–93% by mRNA-1273^25, 30, 39^ and 61%–95% by ChAdOx1^30, 31, 41^). After full vaccination, the VE was all higher than 75% (77%–98% by BNT162b2,^10, 22, 27, 30, 32, 39, 41^ 93%–99% by mRNA-1273,^30, 39^ 79% by ChAdOx1,^41^ 78% by Ad26.COV2.S with single-dose as full vaccination^26^).

**Fig. 2.**
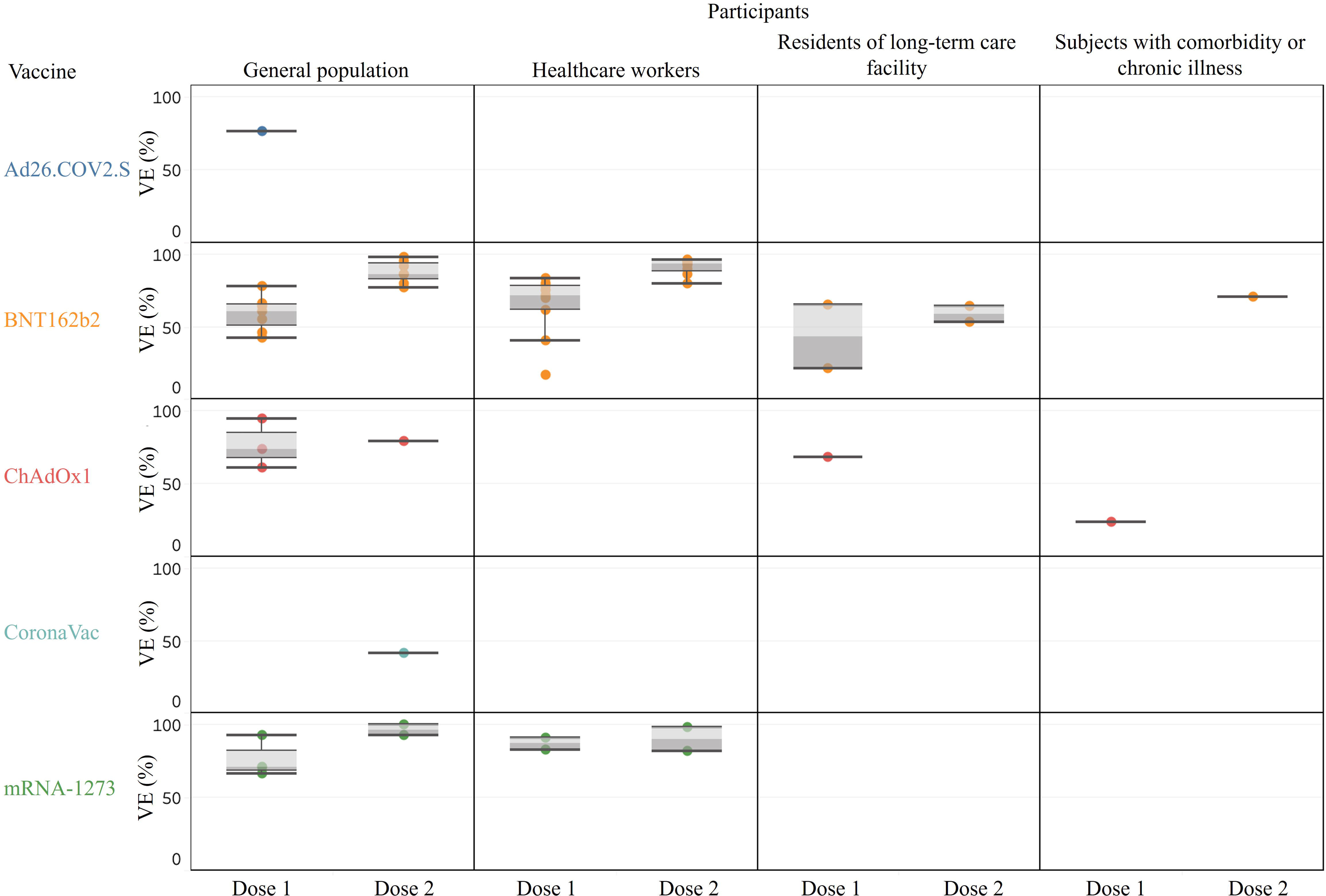
Vaccine effectiveness against overall SARS-CoV-2 infection

The only exception was a low VE noted for CoronaVac (42%, 95% CI, 26.9%–53.3%) in a Brazilian study of subjects aged ≥70, with an 83% of P.1 variants prevalence during study time (Table 1).^42^ The VE among fully vaccinated healthcare workers was 80% or higher (80%–97% by BNT162b2^10, 18, 21, 29, 33, 38, 46, 47^ and 82%–99% by mRNA-1273^46, 47^) (Table 2). Comparatively, VE estimates among residents of long-term care facilities were low (partial vaccinated: 21%–65% by BNT162b2,^38, 44^ 68% [95% CI, 34%–85%] by ChAdOx1;^44^ fully vaccinated: 53%–64% by BNT162b2^10, 38^). And the VE was also relatively low among subjects with comorbidity or chronic illness (one dose of ChAdOx1: 24% after ≥21 days, 50% after ≥42 days,^17^ two doses of BNT162b2: 71% after ≥7 days^10^) (Table 3).

**Table 3.**
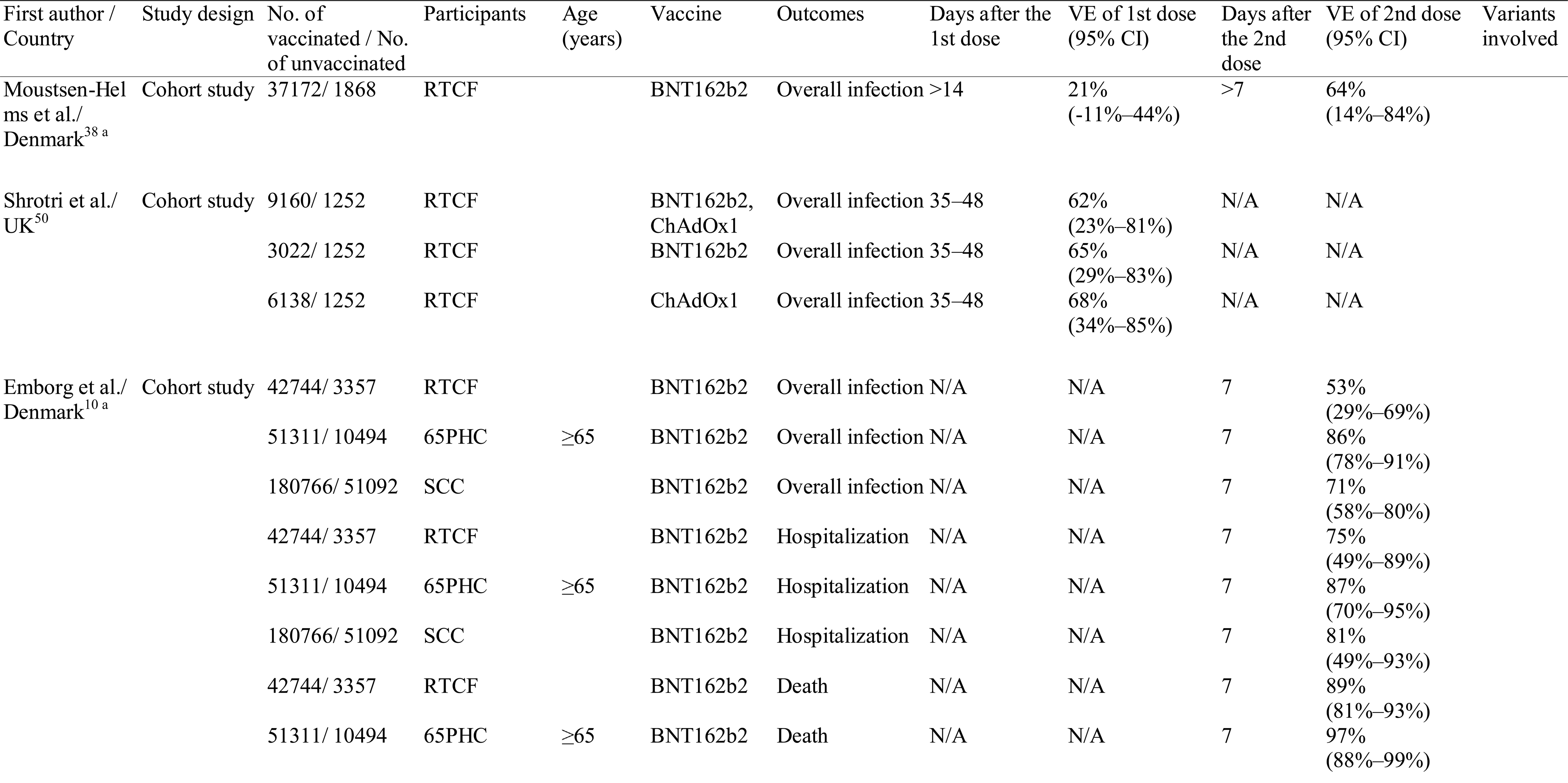
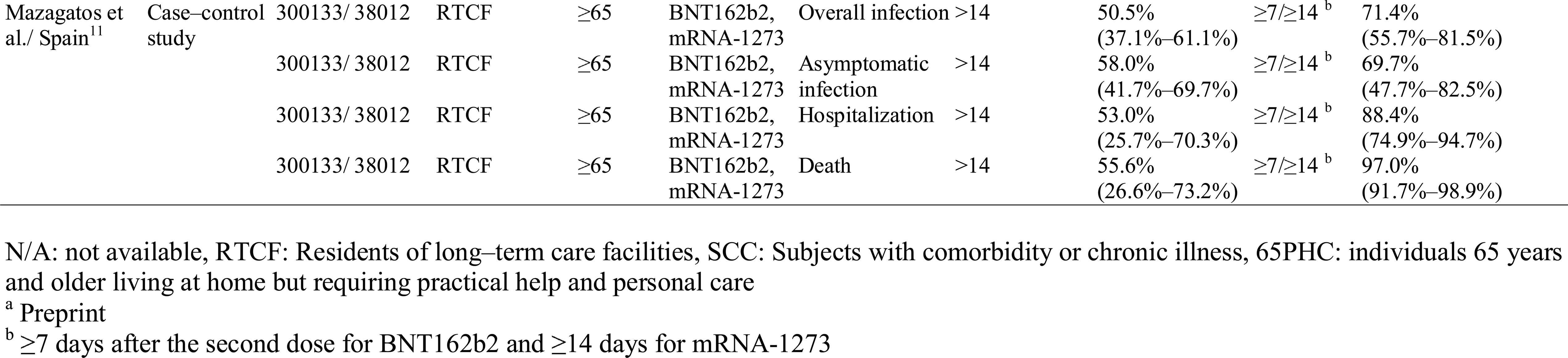
Summary of the studies on the effectiveness of COVID-19 vaccines among residents of long-term care facility, subjects with comorbidity, subjects with chronic illness, or elderly people (≥65 years) requiring personal care

#### Asymptomatic infection, symptomatic infection, hospitalization, critical disease, and death

The VE estimates against SARS-CoV-2 related various outcomes were shown in Table 1–3 and Fig.3–7. Among the general population in the UK, the VE against asymptomatic infection after fully vaccinated either with BNT162b2 (VE: 58%) or ChAdOx1 (VE: 61%) was not as high as that in Israel (VE: 92%) (Fig. 3, Table 1).^32, 41^ Much more beneficial effects were found when symptomatic infection reduction was considered as an outcome, with the VE reaching up to 89%–97% for BNT162b2,^25, 27, 32, 36, 41^ 92% for ChAdOx1,^41^ and 94% for mRNA-1273 after full vaccination (Fig. 4, Table 1).^25^ Studies in healthcare workers also showed beneficial effects from vaccination (VE: 94%–97% by two doses of BNT162b2),^14, 15, 29^ except for a CoronaVac study in Brazil which showed a low VE against symptomatic infection with predominant P.1 variant (37%, 95% CI, 54.9%−74.2%) even after being fully vaccinated (Fig. 4, Table 2).^34^ The protective effects become more obvious for more severe outcomes. The VE against critical disease was higher than 90% by BNT162b2.^27, 32, 39^ A study even demonstrated a VE of 100% for subjects who were fully vaccinated with mRNA-1273 (Fig. 5).^39^

**Fig. 3.**
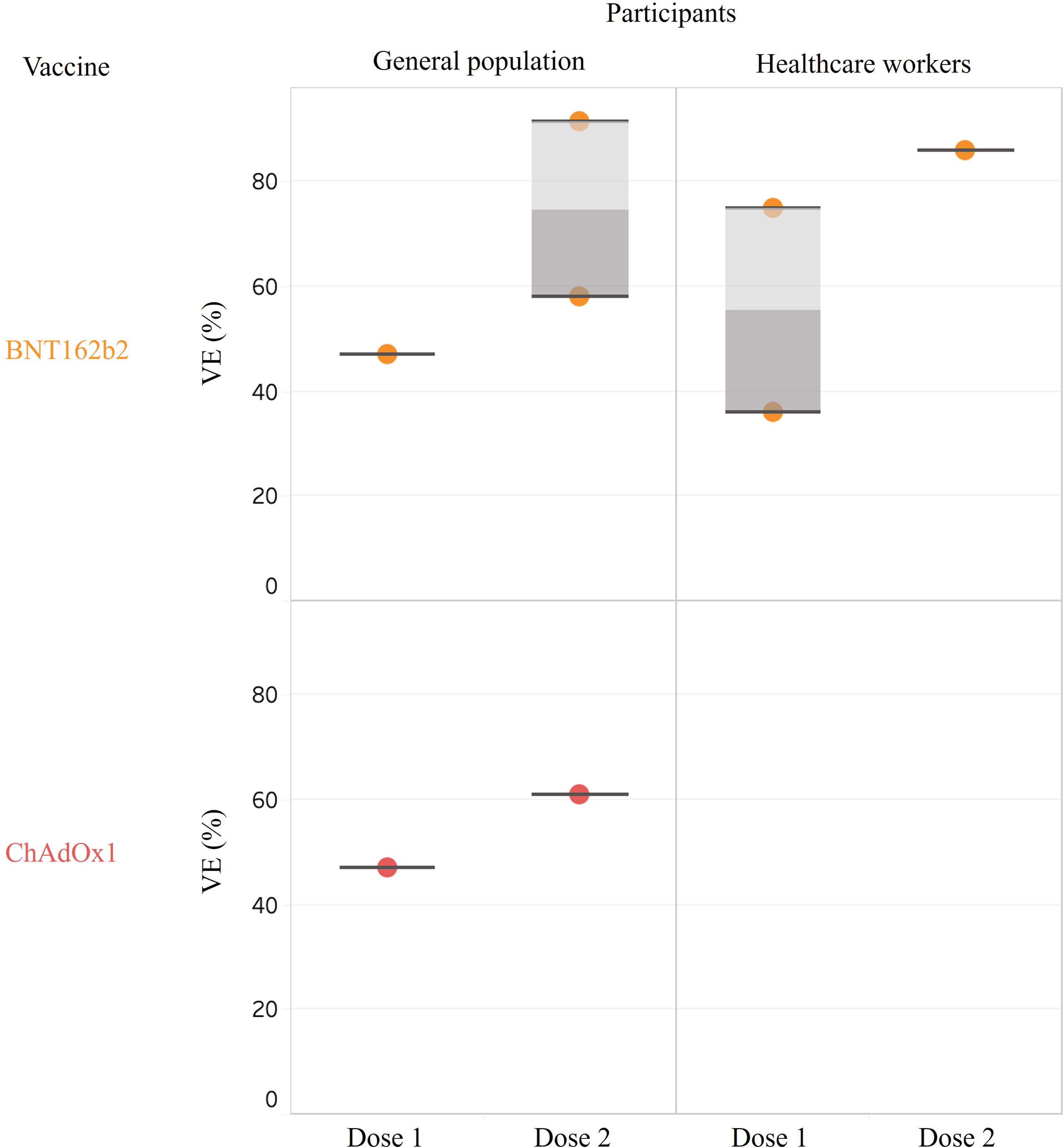
Vaccine effectiveness against asymptomatic infection

**Fig. 4.**
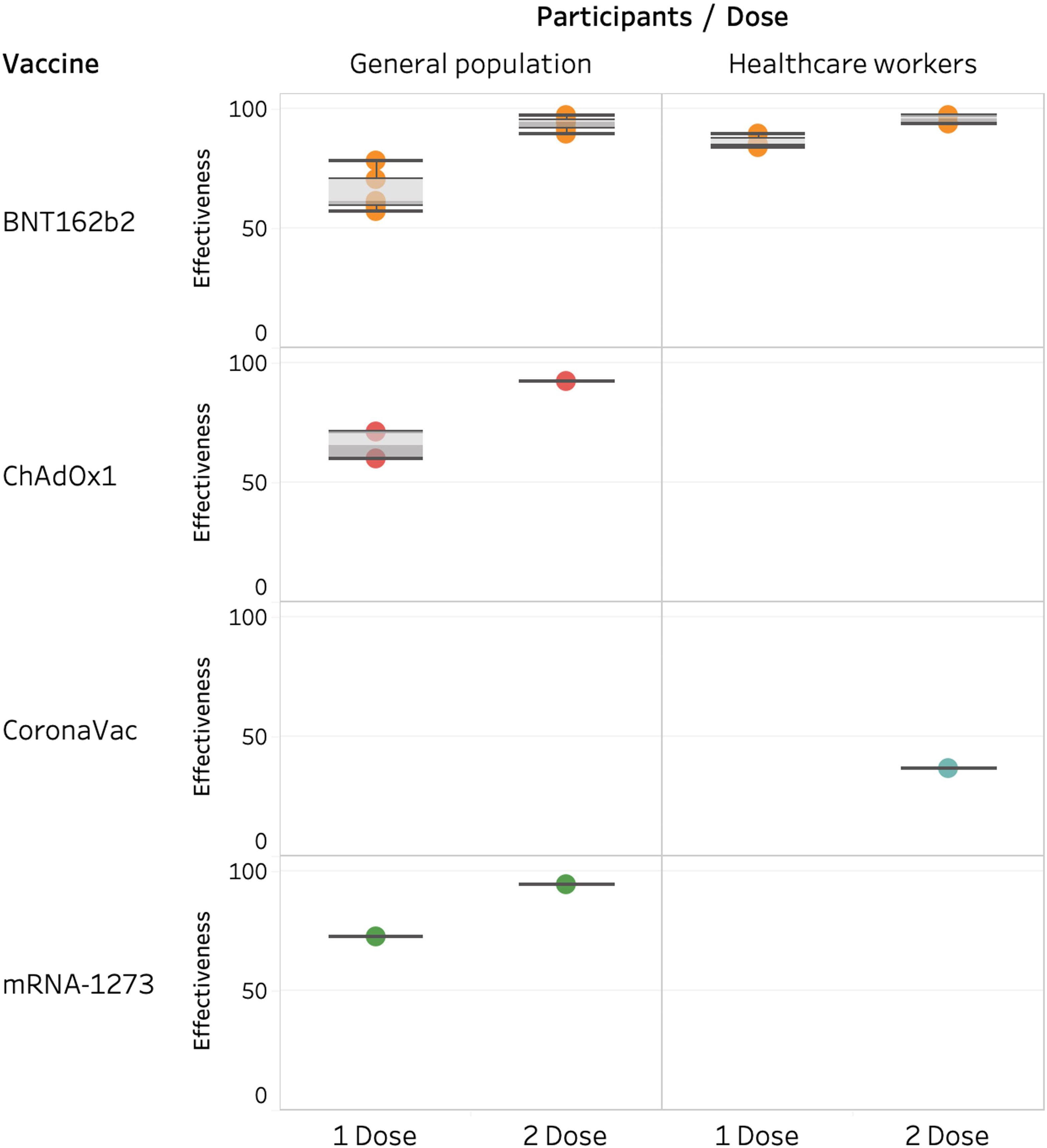
Vaccine effectiveness against symptomatic infection

**Fig. 5.**
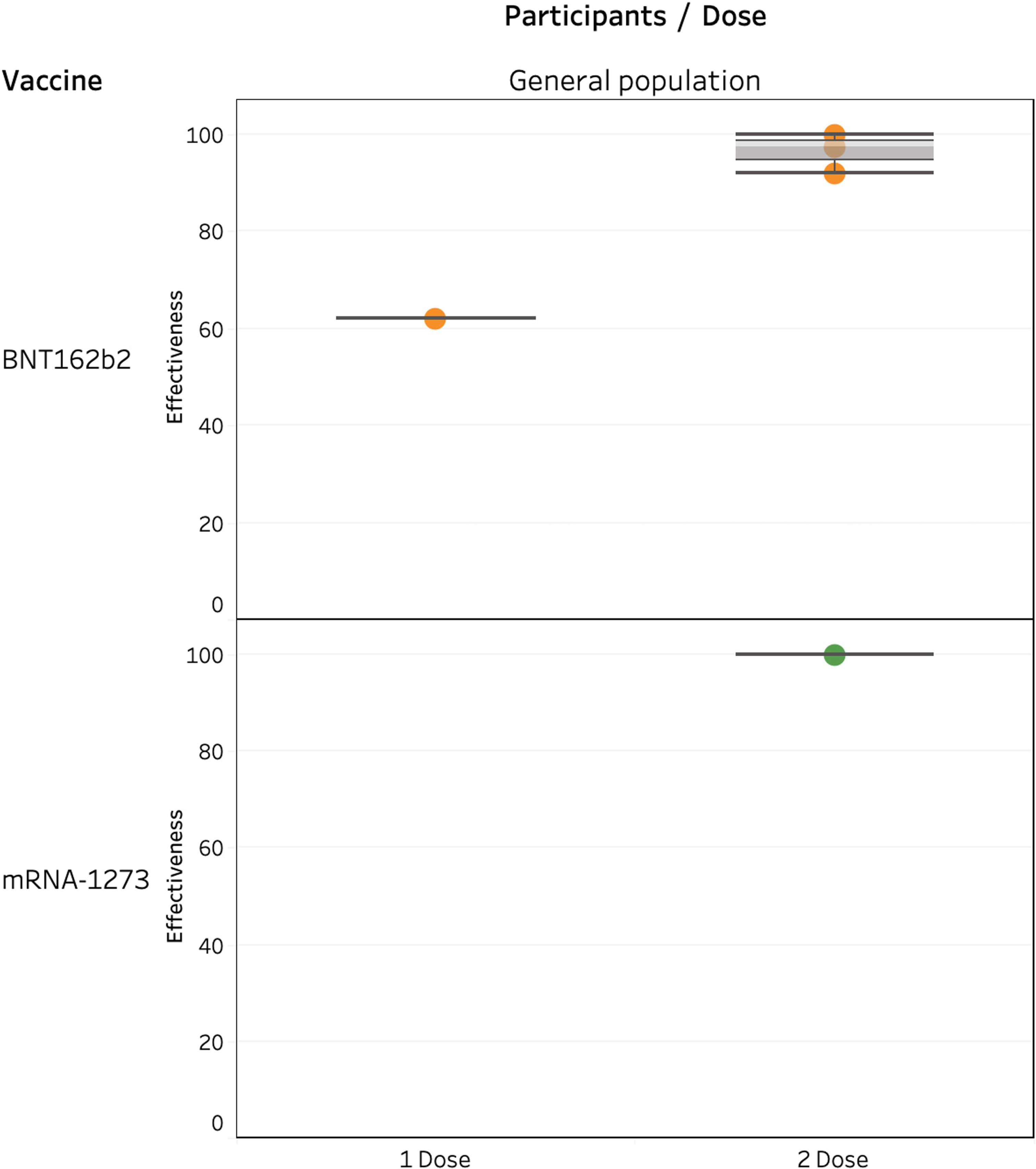
Vaccine effectiveness against critical disease

Regarding the risk reduction in hospitalization among the general population, the effectiveness was similar among one dose of ChAdOx1 (VE: 88%–100%),^30, 49^ two doses of BNT162b2 (VE: 87%–99%),^10, 27, 32, 49^ and two doses of mRNA-1273 (VE: 86%–100%) (Fig. 6).^30, 39^ Risk reduction of death for more than 95% was also found by one dose of ChAdOx1 and 2 doses of mRNA vaccines (Fig. 7). The VE estimates against Covid-19 hospital admissions after vaccination either with BNT162b2 or ChAdOx1 in older age groups were slightly lower than that of younger age groups.^49^ The result of VE across age groups was shown in Supplementary Table S3.

**Fig. 6.**
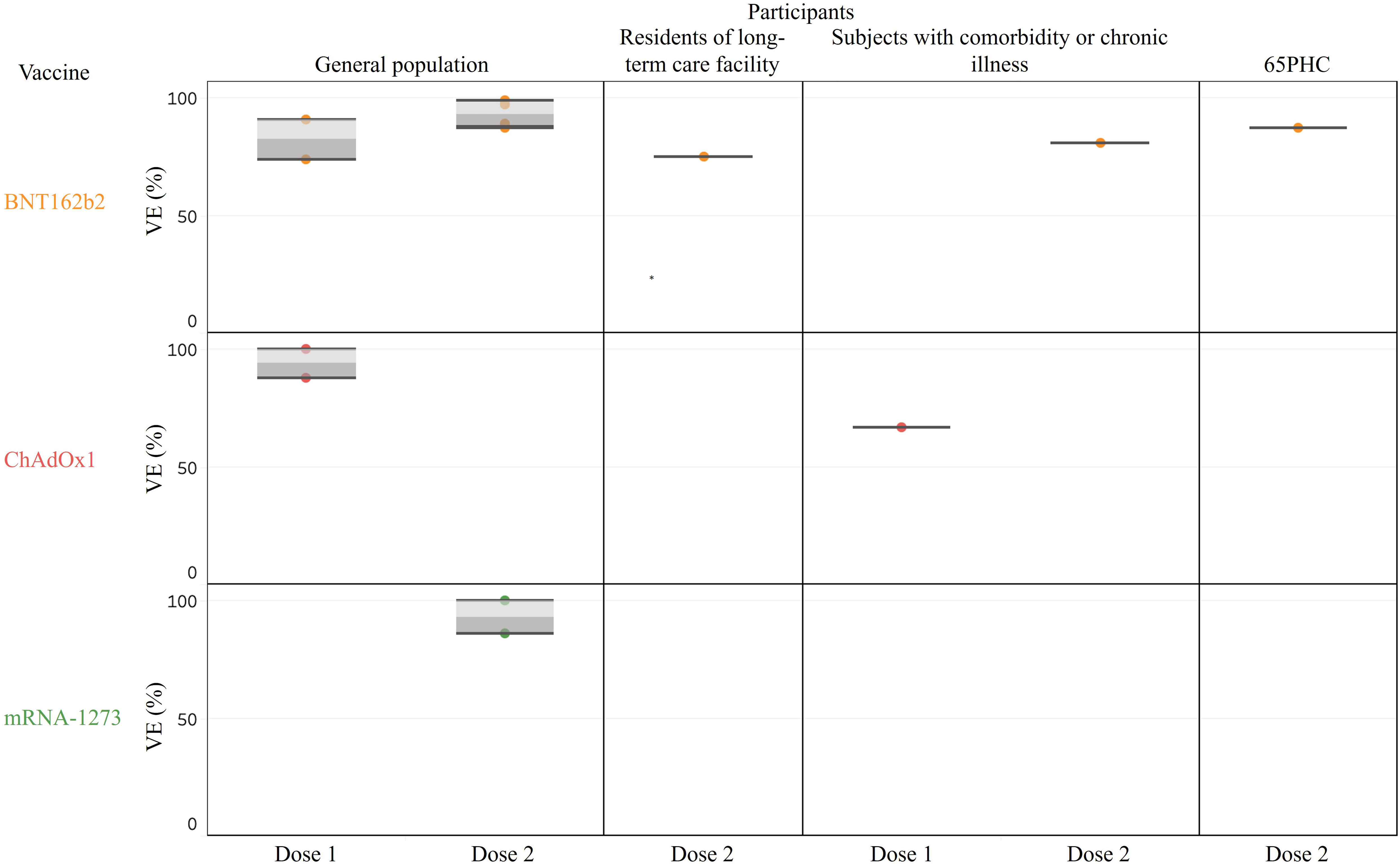
Vaccine effectiveness against hospitalization

**Fig. 7.**
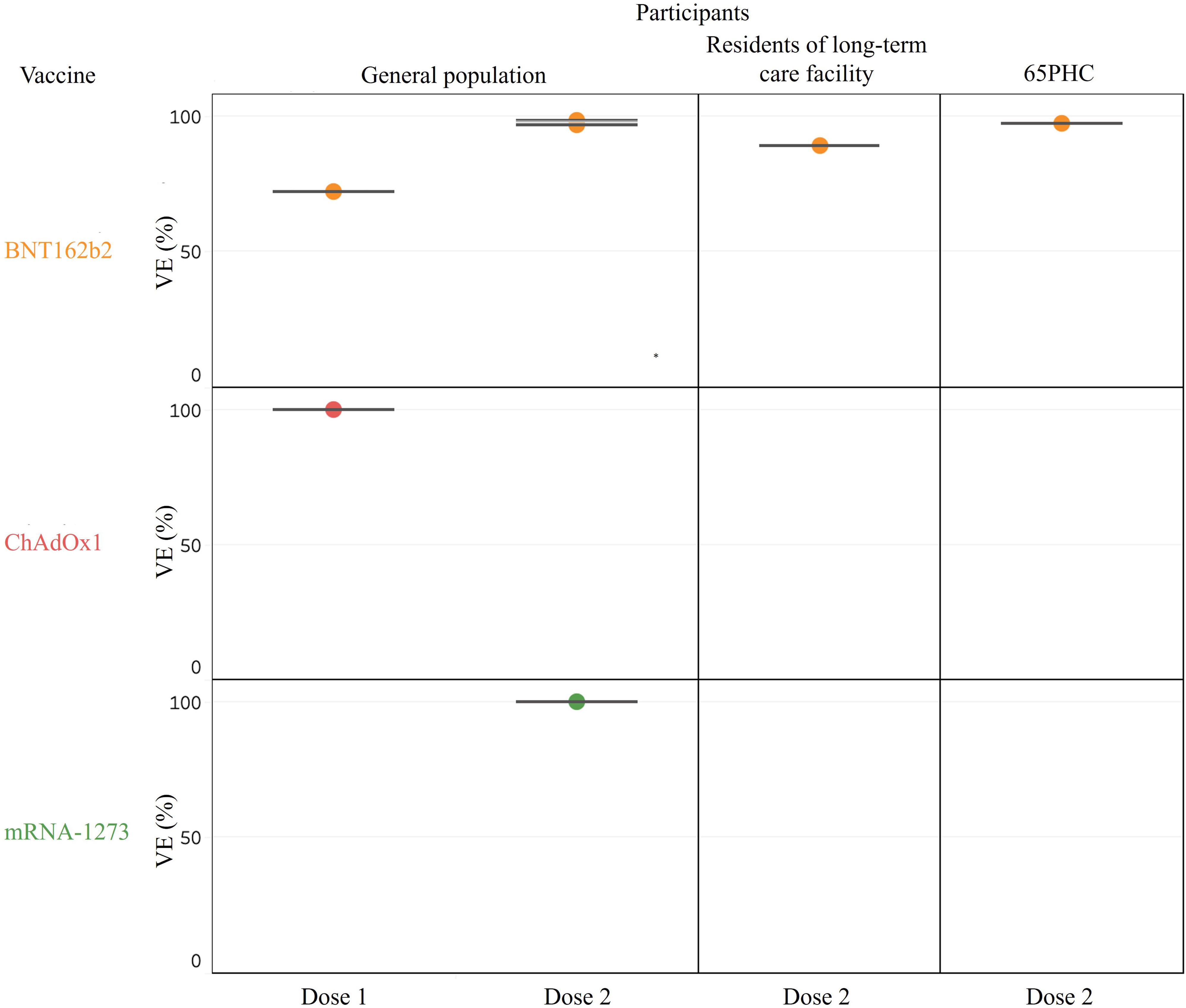
Vaccine effectiveness against death

### Effectiveness of vaccines against SARS-CoV-2 VOC

Three studies have specifically calculated the VE against the VOC ^8, 19, 52, 53^ (Table 1, Fig. 8). For the Alpha variant (B.1.1.7), one dose of BNT162b2 or ChAdOx1 provided around 50% of VE on symptomatic infection and critical disease or death.^8, 53^ For the delta variant (B.1.617.2), better VE was also found in 2 doses of BNT162b2 (VE: 93.4%, [95% CI, 90.4%–95.5%]) than in ChAdOx1 (66.1%, [95% CI, 54.0%–75.0%]) in terms of preventing symptomatic infection.^53^

**Fig. 8.**
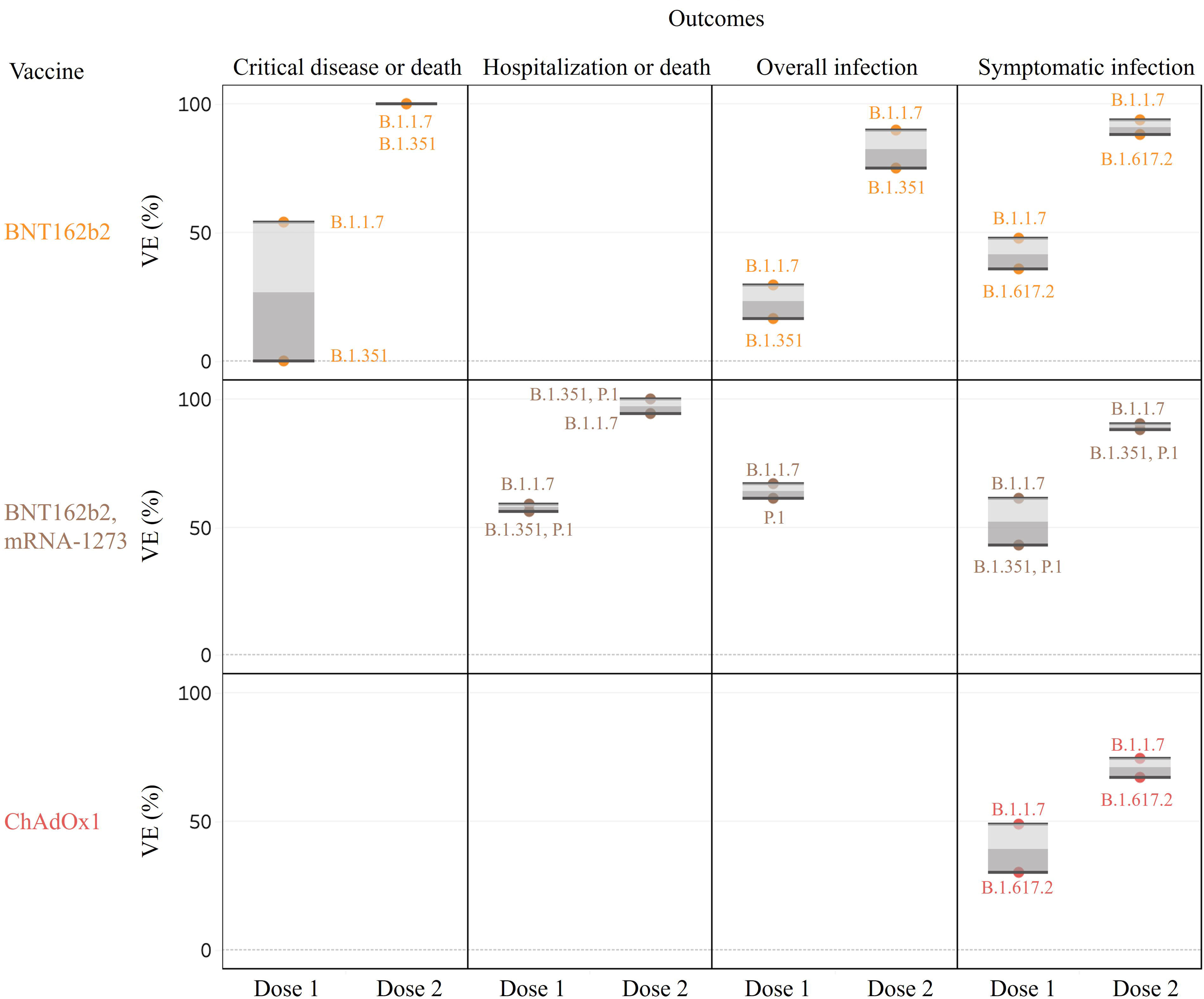
Vaccine effectiveness against SARS-CoV-2 variants of concern

## Discussion

This rapid review included 39 studies from 11 countries. Some of these studies may have overlapping populations with data obtained from the same datasets (e.g. national registry) or from the same institution or same region.^14–16, 18–20, 27, 31, 33, 35–38, 40, 41, 44^ By excluding possible double counted participants, this review approximately covered some 15 million of population in total with over 8 million vaccinated subjects. The results of all included studies are consistently showing that vaccination among the general population in real-world settings, with either BNT162b2, ChAdOx1, or mRNA-1273, substantially reduced the risk of SARS-CoV-2 infection related admission and death. Their protective effectiveness became more obvious as the outcome became more severe. The effectiveness of one dose of ChAdOx1 in preventing hospitalization and death was higher than 80% in general population and similar to that observed in subjects fully vaccinated with BNT162b2 or mRNA-1273. While delta variant is becoming the dominant strain circulating in many parts of the world, VE reports on the delta variant are scarce. One recently published study shows that 2 doses of BNT162b2 can significantly reduce the risk of delta variant-related symptomatic infections.^19^

The suboptimal VE observed in the elderly or residents of long-term care facility are probably due to the immunosenescence which leads to defects in innate and adaptive immune responses. Vaccine responses tended to be weaker and decline earlier ^54^ with a lower titer of antibodies.^55–57^ Since they are vulnerable to developing SARS-CoV-2 infections and its complications, improved vaccine strategies or further vaccine boosting are required in these vulnerable populations.

It should be noted that VE between vaccines cannot be directly compared across these papers due to the following reasons. The first one is the heterogeneity in the interval between vaccination and start of event measurement. The VE of the first dose BNT162b2 against infection was 61% ≥14 days after the vaccination in the study by Pawlowski et al.^39^ and was 72% ≥21 days after vaccination in the study by Hall et al.^33^ For immunity is gradually building,^3^ the protective effects would increase over a given period of time.^17, 27^ At present, there is no data to suggest the timing of the VE plateauing for each vaccine. Second, a large variation is noted regarding the dosing intervals. To vaccinate as many people as possible, the UK government decided that the dosing interval for vaccines (whether BNT162b2 or ChAdOx1) could be extended up to 12 weeks.^20, 36^ In Canada, the administration of the second dose of BNT162b2 and mRNA-1273 was delayed by up to 16 weeks for most individuals due to the disruption of vaccine supply.^25^ There is limited data about the change of the effectiveness for extended dosing intervals. Variations in dosing intervals would be very common in most countries of the world due to the insufficient and unstable vaccine supply. Evidences regarding the effectiveness with appropriate and tolerable dosing intervals are needed.^58, 59^ Third, some studies^11, 28, 37, 40, 43, 48^ estimated the VE by pooled analyses of two vaccines, in which we cannot obtain the VE of each type of vaccines. Fourth, population bias with unmatched control arm or unequal use of vaccine types across study cohorts could make the data of direct comparison of vaccine types unreliable.^19, 31, 36, 39, 49^ Different vaccines may target different populations in the roll-out period. For example, old age and healthcare workers are in the priority group of vaccination program in most of the countries, but the timescale of supply was different between vaccine types.^17, 31^ Although the demographic factors were controlled in most studies, residual confounding may not be entirely excluded.

There are several barriers and limitations in this study. First, literature about the VE against Covid-19 are rapidly evolving and growing exponentially. In this review, 15 of 39 included studies are preprint articles up to June 30, 2021. While we were processing the review and extracting the data in July, five studies have been accepted with their data changed in online version ^19, 31, 34, 37, 45, 51–53, 60^ so that we need to constantly update the data accordingly before submission. Second, these included studies were disproportionately from countries in Europe, North America, and Israel, and were mostly focused on BNT162b2, because these countries have higher priority and access to a large amount of Covid-19 vaccines than most countries in other regions in the first half of 2021.^6^ In addition, shorter dosing intervals and larger vaccine supply in these nations also make the data of VE associated with BNT162b2 much more comprehensive than other vaccines. Longer dosing intervals made the full vaccination reports of ChAdOx1 much fewer in number than those on mRNA vaccines among the included articles over the past months. Furthermore, the VE assessment reports on CoronaVac and Ad26.COV2.S are also too few to draw conclusions. Third, the heterogeneity across studies in dosing interval, timing of outcome measurement, or different target population between vaccine types due to supply chain issues make meta-analyses difficult at present. Fourth, the VE estimates of all the studies were based on relatively short study time (range, <2 months to 6 months) with a median follow-up period ranging from 28 to 106 days.^14, 15, 21, 32, 33, 38, 39, 46^ Further studies are needed to clarify the duration of protective effects of vaccines. The last but not the least important point is the scarce VE reports about the emerging variants such as delta variant which has been currently dominating in the whole world.

## Conclusion

Based on the studies of the first half-year of vaccine administration, this rapid review provided timely and comprehensive evidence on the effectiveness of the ChAdOx1, BNT162b2, and mRNA-1273 vaccines against various Covid-19 infection-related outcomes ranging from asymptomatic to critical illness and death. This review highlights the VE across different segments of populations. These real-world results tended to support the implementation of mass vaccination campaigns as public health strategies, and may also help ease skepticism about VE, which is still a common problem in many parts of the world.

## Supporting information

file:///H:/2021%20COVID%20Vaccine/20210817/Supplementary%20Tables.htm

## Data Availability

The authors confirm that the data supporting the findings of this study are available within the article and its supplementary materials.

