## Supplementary material for "Effectiveness of the WHO-authorized Covid-19 Vaccines: a Rapid Review of Global Reports till June 30, 2021": file:///H:/2021%20COVID%20Vaccine/20210817/Supplementary%20Tables.htm

**Supplementary Tables**

Table S1. Characteristics of the included studies (n=39)

| Country of origin | | |  |
| --- | --- | --- | --- |
|  | The United Kingdom | | 12 |
|  | The United States | | 7 |
|  | Israel | | 7 |
|  | Italy | | 3 |
|  | Denmark | | 2 |
|  | Canada | | 2 |
|  | Brazil | | 2 |
|  | Qatar | | 1 |
|  | Sweden | | 1 |
|  | Spain | | 1 |
|  | Finland | | 1 |
| Study design | | |  |
|  | Cohort | | 30 |
|  | Case control | | 9 |
| Vaccine | | |  |
|  | BNT162b2 | | 36 |
|  | mRNA-1273 | | 12 |
|  | ChAdOx1 | | 10 |
|  | CoronaVac | | 2 |
|  | Ad26.COV2.S | | 1 |
| Study participants | | |  |
|  | | General population | 22 |
|  | | Healthcare workers | 17 |
|  | | Residents of long-term care facility | 4 |
|  | | Individuals with comorbidities or chronic illnesses | 2 |
| Outcomes of interest | | |  |
|  | Infection | | 29 |
|  | Asymptomatic infection | | 5 |
|  | Symptomatic infection | | 13 |
|  | Hospitalization | | 11 |
|  | Severe illness | | 4 |
|  | Death | | 9 |
| SARS-CoV-2 variants of concern included | | |  |
|  | B1.1.7 | | 16 |
|  | B.1.351 | | 2 |
|  | P.1 | | 4 |
|  | B.1.617.2 | | 1 |

Table S2. Risk of bias assessment by ROBINS-I

|  | Bias due to confounding | Bias in selection of participants into the study | Bias in classification of interventions | Bias due to deviations from intended interventions | Bias due to missing data | Bias in measurement of outcomes | Bias in selection of the reported result | Overall bias |
| --- | --- | --- | --- | --- | --- | --- | --- | --- |
| Dagan et al.^27^ | Moderate | Low | Low | Low | Low | Low | Low | Moderate |
| Haas et al.^32^ | Moderate | Low | Low | Low | Low | Low | Low | Moderate |
| Pritchard et al.^41^ | Moderate | Low | Low | Low | Low | Low | Low | Moderate |
| Pawlowski et al.^39^ | Moderate | Low | Low | Low | Low | Low | Low | Moderate |
| Björk et al.^22^ | Serious | Low | Low | Low | Moderate | Low | Low | Serious |
| Abu-Raddad et al.^8^ | Moderate | Low | Low | Low | Low | Low | Low | Moderate |
| Lopez Bernal et al.^61^ | Moderate | Low | Low | Low | Low | Low | Low | Moderate |
| Vasileiou et al.^49^ | Moderate | Low | Low | Low | Low | Low | Low | Moderate |
| Glampson et al.^31, 51^ | Moderate | Low | Low | Low | Low | Low | Low | Moderate |
| Corchado-Garcia et al. ^26^ | Moderate | Low | Low | Low | Low | Low | Low | Moderate |
| Lopez Bernal et al.^20^ | Moderate | Low | Low | Low | Low | Low | Low | Moderate |
| Chung et al.^25^ | Moderate | Low | Low | Low | Low | Low | Low | Moderate |
| Skowronski et al.^45, 52^ | Moderate | Low | Low | Low | Moderate | Low | Low | Moderate |
| Jones et al.^35^ | Moderate | Low | Low | Low | Low | Low | Low | Moderate |
| Fabiani et al.^29^ | Moderate | Low | Low | Low | Low | Low | Low | Moderate |
| Hall et al.^33^ | Moderate | Low | Low | Low | Low | Low | Low | Moderate |
| Pilishvii et al.^40^ | Moderate | Low | Low | Low | Low | Low | Low | Moderate |
| Swift et al.^46^ | Moderate | Low | Low | Low | Low | Low | Low | Moderate |
| Bianchi et al.^21^ | Moderate | Low | Low | Low | Low | Low | Low | Moderate |
| Daniel et al.^28^ | Serious | Serious | Low | Low | Low | Low | Low | Serious |
| Benenson et al.^18^ | Critical | Critical | Low | Low | Low | Low | Low | Critical |
| Amit et al.^14^ | Moderate | Low | Low | Low | Low | Low | Low | Moderate |
| Lumley et al.^37, 60^ | Moderate | Low | Low | Low | Low | Low | Low | Moderate |
| Angel et al.^15^ | Moderate | Low | Low | Low | Low | Low | Low | Moderate |
| Moustsen-Helms et al.^38^ | Moderate | Low | Low | Low | Low | Low | Low | Moderate |
| Shrotri et al.^50^ | Moderate | Low | Low | Low | Low | Low | Low | Moderate |
| Emborg et al.^10^ | Moderate | Low | Low | Low | Low | Low | Low | Moderate |
| Ranzani et al.^42^ | Moderate | Low | Low | Low | Low | Low | Low | Moderate |
| Mazagatos et al.^11^ | Moderate | Low | Low | Low | Low | Low | Low | Moderate |
| Azamgarhi et al.^16^ | Moderate | Low | Low | Low | Low | Low | Low | Moderate |
| Thompson et al.^47^ | Moderate | Low | Low | Low | Moderate | Low | Low | Moderate |
| Lopez Bernal et al.^19^ | Moderate | Low | Low | Low | Low | Low | Low | Moderate |
| Hitchings et al.^34^ | Moderate | Low | Low | Low | Low | Low | Low | Moderate |
| Shrestha et al.^43^ | Moderate | Low | Low | Low | Low | Low | Low | Moderate |
| Vahidy et al.^48^ | Moderate | Low | Low | Low | Low | Low | Low | Moderate |
| Baum et al.^17^ | Moderate | Low | Low | Low | Low | Low | Low | Moderate |
| Chodick et al.^23^ | Moderate | Low | Low | Low | Low | Low | Low | Moderate |
| Chodick et al.^24^ | Moderate | Low | Low | Low | Low | Low | Low | Moderate |
| Flacco et al.^30^ | Moderate | Low | Low | Low | Low | Low | Low | Moderate |

Table S3. Summary of the studies on the effectiveness of COVID-19 vaccines across age groups

| Study first author / Country | Study design | No. of vaccinated / No. of unvaccinated | Participants | Age | Vaccine | Outcome | Days after the 1st dose | VE of 1st dose  (95% CI) | Days after the 2nd dose | VE of 2nd dose  (95% CI) | Variants involved |
| --- | --- | --- | --- | --- | --- | --- | --- | --- | --- | --- | --- |
| Dagan et al./ Israel^27^ | Cohort study | 596618/ 596618 | GP | ≥16 | BNT162b2 | Overall infection | 14–20 | 46%  (40%–51%) | ≥7 | 92%  (88%–95%) | B.1.1.7 |
|  |  | 213090/ 213090 | GP | 16–39 | BNT162b2 | Overall infection | 14–20 | 49%  (41%–57%) | ≥7 | 94%  (87%–93%) | B.1.1.7 |
|  |  | 304514/ 304514 | GP | 40–69 | BNT162b2 | Overall infection | 14–20 | 47%  (40%–55%) | ≥7 | 90%  (82%–95%) | B.1.1.7 |
|  |  | 79014/ 79014 | GP | ≥70 | BNT162b2 | Overall infection | 14–20 | 22%  (-9%–44%) | ≥7 | 95%  (87%–100%) | B.1.1.7 |
|  |  | 596618/ 596618 | GP | ≥16 | BNT162b2 | Symptomatic infection | 14–20 | 57%  (50%–63%) | ≥7 | 94%  (87%–98%) | B.1.1.7 |
|  |  | 213090/ 213090 | GP | 16–39 | BNT162b2 | Symptomatic infection | 14–20 | 57%  (46%–68%) | ≥7 | 99%  (96%–100%) | B.1.1.7 |
|  |  | 304514/ 304514 | GP | 40–69 | BNT162b2 | Symptomatic infection | 14–20 | 59%  (50%–67%) | ≥7 | 90%  (75%–98%) | B.1.1.7 |
|  |  | 79014/ 79014 | GP | ≥70 | BNT162b2 | Symptomatic infection | 14–20 | 44%  (19%–64%) | ≥7 | 98%  (90%–100%) | B.1.1.7 |
| Haas et al./ Israel^32^ | Cohort study | 4714932/ 1823979 ^a^ | GP | ≥16 | BNT162b2 | Overall infection | N/A | N/A | ≥7 | 95.3%  (94.9%–95.7%) | B.1.1.7 |
|  |  | 2290820/ 1356028 ^a^ | GP | 16–44 | BNT162b2 | Overall infection | N/A | N/A | ≥7 | 96.1%  (95.7%–96.5%) | B.1.1.7 |
|  |  | 1408492/ 355606 ^a^ | GP | 45–64 | BNT162b2 | Overall infection | N/A | N/A | ≥7 | 94.9%  (94.2%–95.5%) | B.1.1.7 |
|  |  | 1015620/ 112345 ^a^ | GP | ≥65 | BNT162b2 | Overall infection | N/A | N/A | ≥7 | 94.8%  (93.9%–95.5%) | B.1.1.7 |
|  |  | 4714932/ 1823979 ^a^ | GP | ≥16 | BNT162b2 | Asymptomatic infection | N/A | N/A | ≥7 | 91.5%  (90.7%–92.2%) | B.1.1.7 |
|  |  | 2290820/ 1356028 ^a^ | GP | 16–44 | BNT162b2 | Asymptomatic infection | N/A | N/A | ≥7 | 93.6%  (92.8%–94.4%) | B.1.1.7 |
|  |  | 1408492/ 355606 ^a^ | GP | 45–64 | BNT162b2 | Asymptomatic infection | N/A | N/A | ≥7 | 90.8%  (89.6%–91.9%) | B.1.1.7 |
|  |  | 1015620/ 112345 ^a^ | GP | ≥65 | BNT162b2 | Asymptomatic infection | N/A | N/A | ≥7 | 88.5%  (86.4%–90.3%) | B.1.1.7 |
|  |  | 4714932/ 1823979 ^a^ | GP | ≥16 | BNT162b2 | Symptomatic infection | N/A | N/A | ≥7 | 97.0%  (96.7%–97.2%) | B.1.1.7 |
|  |  | 2290820/ 1356028 ^a^ | GP | 16–44 | BNT162b2 | Symptomatic infection | N/A | N/A | ≥7 | 97.6%  (97.3%–97.8%) | B.1.1.7 |
|  |  | 1408492/ 355606 ^a^ | GP | 45–64 | BNT162b2 | Symptomatic infection | N/A | N/A | ≥7 | 96.7%  (96.3%–97.0%) | B.1.1.7 |
|  |  | 1015620/ 112345 ^a^ | GP | ≥65 | BNT162b2 | Symptomatic infection | N/A | N/A | ≥7 | 96.4%  (95.9%–97.0%) | B.1.1.7 |
|  |  | 4714932/ 1823979 ^a^ | GP | ≥16 | BNT162b2 | Hospitalization | N/A | N/A | ≥7 | 97.2%  (96.8%–97.5%) | B.1.1.7 |
|  |  | 2290820/ 1356028 ^a^ | GP | 16–44 | BNT162b2 | Hospitalization | N/A | N/A | ≥7 | 98.1%  (97.3%–98.7%) | B.1.1.7 |
|  |  | 1408492/ 355606 ^a^ | GP | 45–64 | BNT162b2 | Hospitalization | N/A | N/A | ≥7 | 97.6%  (97.1%–98.1%) | B.1.1.7 |
|  |  | 1015620/ 112345 ^a^ | GP | ≥65 | BNT162b2 | Hospitalization | N/A | N/A | ≥7 | 96.8%  (96.2%–97.3%) | B.1.1.7 |
|  |  | 4714932/ 1823979 ^a^ | GP | ≥16 | BNT162b2 | Critical disease | N/A | N/A | ≥7 | 97.5%  (97.1%–97.8%) | B.1.1.7 |
|  |  | 2290820/ 1356028 ^a^ | GP | 16–44 | BNT162b2 | Critical disease | N/A | N/A | ≥7 | 98.9%  (97.6%–99.5%) | B.1.1.7 |
|  |  | 1408492/ 355606 ^a^ | GP | 45–64 | BNT162b2 | Critical disease | N/A | N/A | ≥7 | 98.1%  (97.5%–98.5%) | B.1.1.7 |
|  |  | 1015620/ 112345 ^a^ | GP | ≥65 | BNT162b2 | Critical disease | N/A | N/A | ≥7 | 97.3%  (96.8%–97.8%) | B.1.1.7 |
|  |  | 4714932/ 1823979 ^a^ | GP | ≥16 | BNT162b2 | Death | N/A | N/A | ≥7 | 96.7%  (96.0%–97.3%) | B.1.1.7 |
|  |  | 2290820/ 1356028 ^a^ | GP | 16–44 | BNT162b2 | Death | N/A | N/A | ≥7 | 100% | B.1.1.7 |
|  |  | 1408492/ 355606 ^a^ | GP | 45–64 | BNT162b2 | Death | N/A | N/A | ≥7 | 95.8%  (92.6%–97.6%) | B.1.1.7 |
|  |  | 1015620/ 112345 ^a^ | GP | ≥65 | BNT162b2 | Death | N/A | N/A | ≥7 | 96.9%  (96.0%–97.6%) | B.1.1.7 |
| Pritchard et al./ UK^41^ | Case–control study | 67738/ 192224 | GP | ≥16 | BNT162b2 | Overall infection | ≥ 21 | 66%  (60%–71%) | ≥1 | 80%  (73%–85%) | B.1.1.7 |
|  |  | 123850/ 192224 | GP | ≥16 | ChAdOx1 | Overall infection | ≥ 21 | 61%  (54%–68%) | ≥1 | 79%  (65%–88%) | B.1.1.7 |
|  |  |  | GP | <75 | BNT162b2, ChAdOx1 | Overall infection | ≥ 21 | 60%  (54%–65%) | N/A | N/A | B.1.1.7 |
|  |  |  | GP | ≥75 | BNT162b2, ChAdOx1 | Overall infection | ≥ 21 | 72%  (64%–78%) | N/A | N/A | B.1.1.7 |
| Vasileiou et al./ UK^49^ | Cohort study | 1331993/ 3077595 | GP | ≥18 | BNT162b2, ChAdOx1 | Hospitalization | 28–34 | 89%  (83%–92%) | N/A | N/A |  |
|  |  | 711839/ 3077595 | GP | ≥18 | BNT162b2 | Hospitalization | 28–34 | 91%  (85%–94%) | N/A | N/A |  |
|  |  | 620154/ 3077595 | GP | ≥18 | ChAdOx1 | Hospitalization | 28–34 | 88%  (75%–94%) | N/A | N/A |  |
|  |  | 470960/ 2913484 | GP | 18–64 | BNT162b2, ChAdOx1 | Hospitalization | 28–34 | 92%  (82%–97%) | N/A | N/A |  |
|  |  | 651924/ 107022 | GP | 65–79 | BNT162b2, ChAdOx1 | Hospitalization | 28–34 | 93%  (73%–98%) | N/A | N/A |  |
|  |  | 209109/ 57089 | GP | ≥80 | BNT162b2, ChAdOx1 | Hospitalization | 28–34 | 83%  (72%–89%) | N/A | N/A |  |
|  |  | 359434/ 2913484 | GP | 18–64 | BNT162b2 | Hospitalization | 28–34 | 92%  (82%–97%) | N/A | N/A |  |
|  |  | 315620/ 107022 | GP | 65–79 | BNT162b2 | Hospitalization | 28–34 | 93%  (73%–98%) | N/A | N/A |  |
|  |  | 36785/ 57089 | GP | ≥80 | BNT162b2 | Hospitalization | 28–34 | 88%  (76%–94%) | N/A | N/A |  |
|  |  | 111526/ 2913484 | GP | 18–64 | ChAdOx1 | Hospitalization | 28–34 | 100%  (NA–100%) | N/A | N/A |  |
|  |  | 336304/ 107022 | GP | 65–79 | ChAdOx1 | Hospitalization | 28–34 | 100%  (NA–100%) | N/A | N/A |  |
|  |  | 172324/ 57089 | GP | ≥80 | ChAdOx1 | Hospitalization | 28–34 | 81%  (60%–91%) | N/A | N/A |  |
| Ranzani et al./ Brazil^42^ ^b^ | Case–control study | 4854/ 11046 | GP | ≥70 | CoronaVac | Overall infection | N/A | N/A | ≥14 | 41.6%  (26.9%–53.3%) | P.1 |
|  |  |  | GP | 70–74 | CoronaVac | Overall infection | N/A | N/A | ≥14 | 61.8%  (34.8%–77.7%) | P.1 |
|  |  |  | GP | 75–79 | CoronaVac | Overall infection | N/A | N/A | ≥14 | 48.9%  (23.3%–66.0%) | P.1 |
|  |  |  | GP | ≥80 | CoronaVac | Overall infection | N/A | N/A | ≥14 | 28.0%  (0.6%–47.9%) | P.1 |
| Chodick et al./ Israel^23^ | Cohort study | 503875  (351897 had follow–up data for days 13 to 24) | GP | ≥16 | BNT162b2 | Overall infection | 13–24 | 51.4%  (16.3%–71.8%) | N/A | N/A |  |
|  |  |  | GP | <60 | BNT162b2 | Overall infection | 13–24 | 50.2%  (14.1%–71.2%) | N/A | N/A |  |
|  |  |  | GP | ≥60 | BNT162b2 | Overall infection | 13–24 | 44.5%  (4.2%–67.9%) | N/A | N/A |  |
| Chodick et al./ Israel^24^ | Cohort study | 1178597  (872454 reach protection period) | GP | ≥16 | BNT162b2 | Overall infection | N/A | N/A | 7–27 | 90%  (79%–95%) |  |
|  |  |  | GP | 16–44 | BNT162b2 | Overall infection | N/A | N/A | 7–27 | 92%  (83%–96%) |  |
|  |  |  | GP | 45–64 | BNT162b2 | Overall infection | N/A | N/A | 7–27 | 90%  (80%–95%) |  |
|  |  |  | GP | 65–74 | BNT162b2 | Overall infection | N/A | N/A | 7–27 | 82%  (63%–92%) |  |
|  |  |  | GP | ≥75 | BNT162b2 | Overall infection | N/A | N/A | 7–27 | 82%  (61%–91%) |  |

GP: general population, N/A: not available

^a^ No. of fully vaccinated / No. of not fully vaccinated

^b^ Preprint
